# Integrative genomic and epigenomic profiling in plasma and urinary cell-free DNA improves early risk stratification of newly diagnosed prostate cancer

**DOI:** 10.1101/2025.04.13.25325732

**Authors:** Anja Lisa Riediger, Samaneh Eickelschulte, Florian Janke, Daniela Janscho, Olga Lazareva, Daniel Hübschmann, Stefan Duensing, Oliver Stegle, Holger Sültmann, Magdalena Görtz

**Author notes:** Corresponding author(s):* Anja Lisa Riediger Im Neuenheimer Feld 460, 69120 Heidelberg, Germany Magdalena Görtz Im Neuenheimer Feld 223, 69120 Heidelberg, Germany.

## Abstract

**Background and Objective:** Prostate cancer (PCa) is a heterogeneous disease, impeding early detection and risk stratification. Liquid biopsies (LBx) enable minimally invasive tumor profiling, but circulating tumor-derived DNA (ctDNA) detection remains difficult, especially in early-stage PCa. This study aimed at developing a multimodal LBx approach, analyzing genomic and epigenomic cell-free DNA (cfDNA) features in plasma and urine from newly diagnosed PCa patients for early detection, tumor characterization, and risk stratification of aggressive PCa.

**Methods:** Plasma and urine samples were included from 55 localized PCa (lPCa) patients, 18 advanced PCa (aPCa) patients, and 36 cancer-free controls. Low-coverage whole-genome sequencing and methylated DNA immunoprecipitation sequencing were performed to assess fragmentation, chromosomal instability, and methylation in cfDNA.

**Key findings and Limitations:** The complementary (epi)genomic analysis of plasma and urinary cfDNA achieved a 45% ctDNA detection rate in newly diagnosed PCa. Major differences were observed between aPCa and controls, reflecting increasing signals with tumor progression. Epigenomic cfDNA features differentiated lPCa from aPCa, and ctDNA was detected in 46% of PCa patients with prostate-specific antigen <10 ng/ml, suggesting potential for risk stratification. However, sensitivity in early PCa remains a major limitation.

**Conclusions and Clinical Implications:** This study highlights the potential of multimodal LBx approaches, integrating genomic and epigenomic cfDNA features, for minimally invasive characterization of primary PCa and potential metastasis at initial diagnosis. While promising for risk stratification, sensitivity requires optimization for early detection. Incorporating LBx into clinical workflows could complement diagnostics and support clinical decision-making for personalized treatments tailored to patients’ PCa risk profiles.

## 1 Introduction

Prostate cancer (PCa) is a clinically and molecularly heterogeneous disease, impeding risk stratification and treatment decisions^1,2^. Current diagnostics provide limited insight into tumor heterogeneity^3^, highlighting the need for additional biomarkers to enable more precise, patient-tailored management. Liquid biopsy (LBx) facilitates molecular characterization of the tumor and its metastasis, allowing for non-invasive monitoring of tumor progression^4,5^. The analysis of cell-free DNA (cfDNA) is a well-established approach for the detection of genomic alterations, indicating the presence of circulating tumor DNA (ctDNA) and enabling tumor burden estimation^6^. CtDNA detection in PCa is challenging, due to overall limited ctDNA shedding and mutational burden^7,8^. Epigenomic profiling holds promise for early tumor characterization, given the early occurrence and tissue specificity of epigenomic changes, such as DNA methylation^9,10^. CfDNA fragmentation patterns were equally shown to harbor tumor-specific information and reflect ctDNA levels^11–13^. This study developed a multimodal LBx approach, analyzing (epi)genomic cfDNA features with low-coverage whole-genome sequencing (lcWGS) and cell-free methylated DNA immunoprecipitation sequencing (cfMeDIP-seq) in plasma and urine of newly diagnosed PCa patients and individuals without cancer. The complementary analysis aimed to improve ctDNA detection, supporting tumor characterization and early risk stratification.

## 2 Materials and Methods

The Supplementary Material provides detailed descriptions of all method sections.

### Study patients

Seventy-three PCa patients at initial diagnosis and 36 cancer-free controls were recruited at Heidelberg University Hospital (June 2021 – November 2022). The control cohort included men undergoing PCa screening or treatment for benign urological conditions. Blood and urine samples were collected prior to examinations, i.e., prostate biopsy or surgery. All individuals provided informed consent; the study was approved by the ethic committee of the Medical Faculty of Heidelberg University (S-130/2021).

### Sample preparation and sequencing

Peripheral blood and urine were processed by double-spin centrifugation within 6 hours. CfDNA was isolated from 1–5.5 ml of plasma and 9.5–19 ml of urine supernatant, respectively, using the QIAamp MinElute ccfDNA Kit (Qiagen). Eight fresh-frozen PCa tissue samples were provided by the Tissue Bank of the National Center of Tumor Diseases (NCT) Heidelberg. Genomic DNA (gDNA) from PCa tissue and matched buffy coat was extracted using the AllPrep DNA/RNA/Protein Mini Kit (Qiagen). Libraries were prepared from 2.4–7 ng cfDNA or 100 ng gDNA, using the KAPA HyperPrep Kit (Roche) with NEBNext UDI-UMI Adaptors (New England Biolabs), followed by a methylated DNA immunoprecipitation workflow^14^. One part of the volume was not enriched. Enriched and non-enriched libraries were amplified for 12-13 (gDNA: eight) and 8–9 (gDNA: six) cycles of polymerase chain reaction, respectively, pooled equimolarly, and paired-end sequenced (2×100 bp) on the NovaSeq 6000 platform (Illumina).

### Sequencing data analyses

Raw sequencing data was processed with a custom pipeline, including adapter trimming, alignment to human genome (hg19), deduplication, and quality filtering. LcWGS data (non-enriched libraries) was applied for cfDNA fragmentation analysis, chromosomal instability analysis (CIA), and for copy number profiling with tumorfraction (TFx) estimation using the ichorCNA algorithm^15^. Genome-wide methylation profiling was performed based on the (cf)MeDIP-seq data, using the R package MESA v0.2.2^16^. Differentially methylated regions (DMRs) were assessed in plasma and urine between tumor patients and controls. DMRs were identified in PCa tissue relative to matched buffy coat, and validated using a published dataset^17^ to determine PCa-specific methylation markers. A synoptic methylation score was calculated for LBx samples based on these markers. The following (epi)genomic features were applied for ctDNA detection: TFx, CIA score, methylation score, 10bp-oscillation score (plasma) and P163–169bp (urine).

### Statistical analyses and data visualization

Statistical analyses were conducted in R v4.0.0^18^; plots were generated using R package ggplot2^19^. Statistical significance was assessed using the Kruskal-Wallis test with Dunn’s post hoc test, Wilcoxon rank-sum test, Fisher’s exact test, and Spearman’s correlation. P values were adjusted with Benjamini-Hochberg‘s method. Cumulative frequency distributions of cfDNA fragmentation were compared with Kolmogorov-Smirnov testing. Significance was set at (adjusted) p < 0.05, unless stated otherwise. The ctDNA detectability threshold for all evaluated (epi)genomic markers was defined as >95^th^ percentile (10bp-oscillation score: <5^th^ percentile) of the control cohort.

## 3 Results

### Patient characteristics

Seventy-three patients with newly diagnosed PCa and 36 cancer-free controls were enrolled (**Table 1**). The majority (75%) had localized disease, while 25% presented with lymph node or distant metastases. LcWGS and (cf)MeDIP-seq were performed on 109 plasma and 102 urine samples (**Table 1**), as well as on PCa tissue and matched buffy coat samples from eight PCa patients.

**Table 1:**
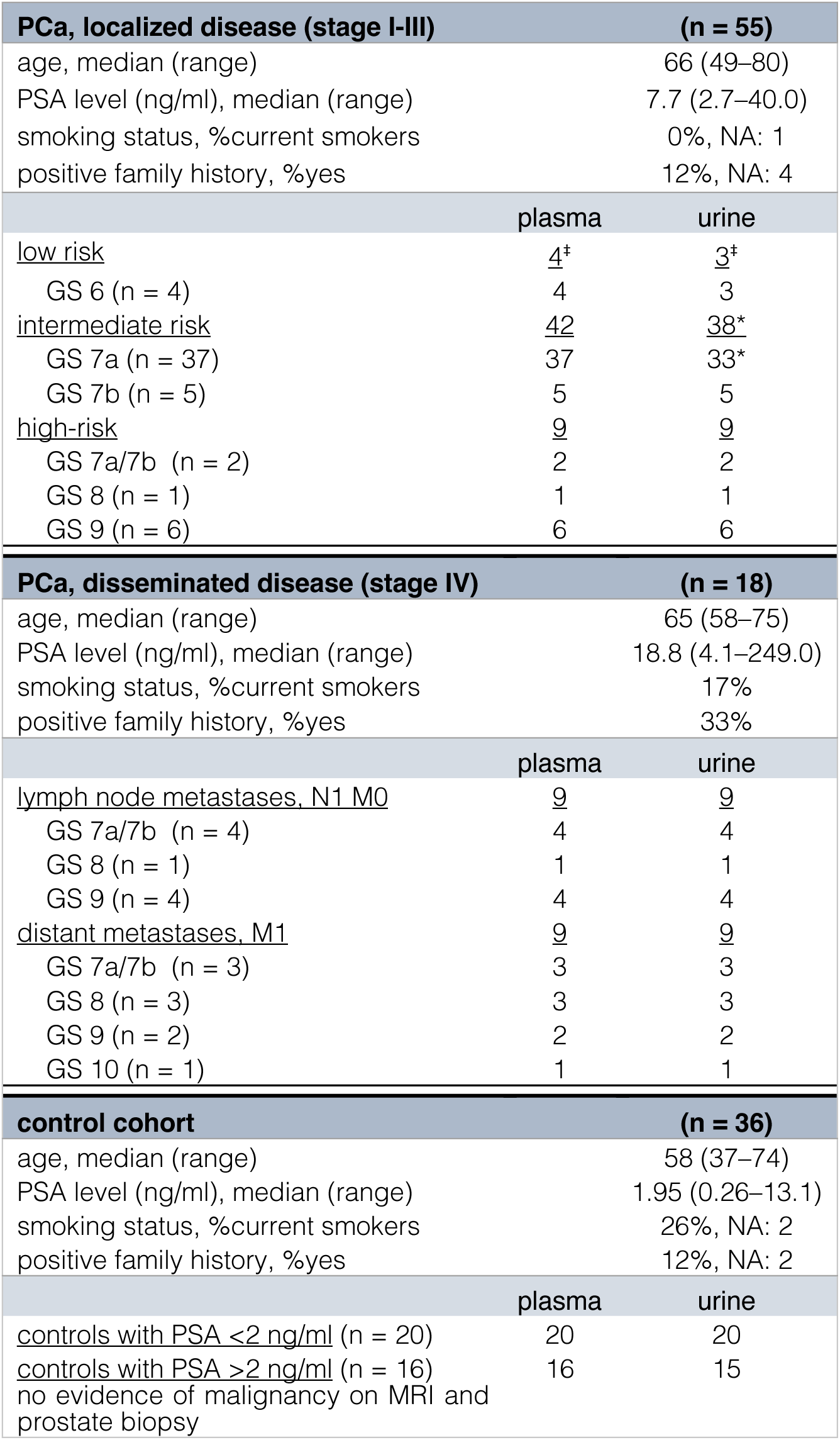
Patients and LBx sample overview. PCa stages were determined based on UICC (stage I-IV) ^39^ and D’Amico Risk Classification^40^. LBx = Liquid Biopsy, M0/M1 = presence/absence of distant metastases, MRI = magnetic resonance imaging, N1 = presence of lymph node metastases, GS = Gleason Score, NA = data not available, n = number, PCa = prostate cancer, PSA = prostate-specific antigen, UICC = Union for International Cancer control, ^‡^ one patient had additional sample collection after one year with upgrade to PCa GS 7b, *one sample excluded after sequencing quality control

### CfDNA methylation patterns distinguished metastatic from non-metastatic PCa and controls

Genome-wide methylation profiling in LBx was conducted to detect tumor-informative regions with aberrant cfDNA methylation. Differential analysis revealed no significant DMRs between all PCa samples and controls (*Supplementary Table S2*). However, metastatic PCa (mPCa) samples showed distinct methylation patterns in both plasma and urine compared to controls and non-mPCa. In plasma cfDNA, 712 DMRs were identified in mPCa compared to controls, and 890 DMRs in mPCa vs. non-mPCa (**Fig. 1A**). Overlapping these sets revealed 445 shared DMRs, including 392 hypermethylated and 21 hypomethylated regions in mPCa (*Supplementary Table S2 + Figure S1)*. In urinary cfDNA, only few DMRs were identified: 48 in mPCa vs. controls and 64 in mPCa vs. non-mPCa, with 16 overlapping regions (13x hypermethylated, 3x hypomethylated; **Fig. 1A**; *Supplementary Figure S1*). Comparison of DMRs in plasma and urine revealed eight shared regions in mPCa vs. controls, and seven in mPCa vs. non-mPCa, including three identical hypermethylated regions, associated with the genes *SOX2-OT*, *ZBTB46*, and *PTPRN2* (**Fig. 1A**).

**Fig. 1:**
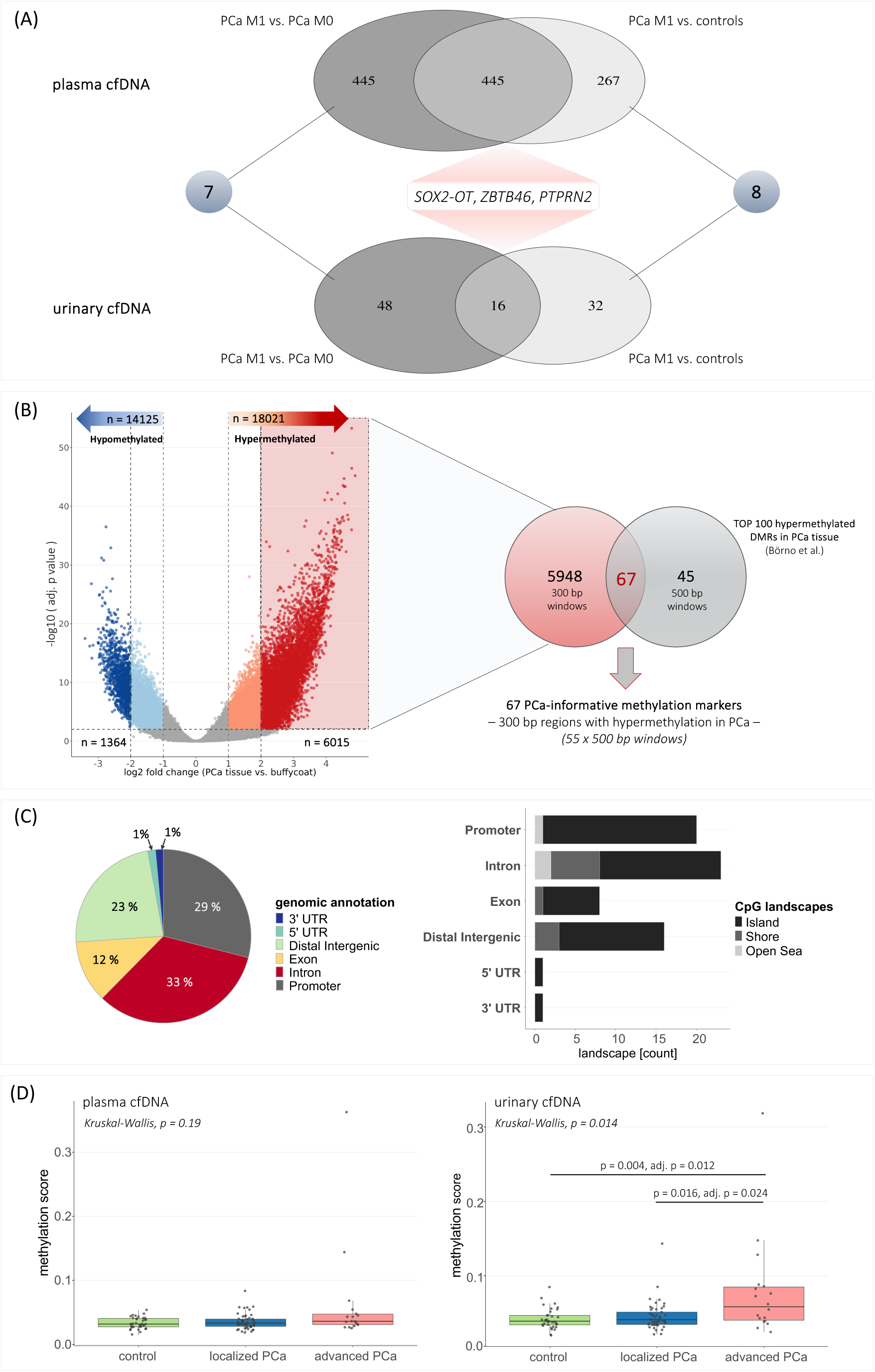
Genome-wide methylation profiling in PCa tissue and LBx samples. (A) Results from the determination of DMRs between mPCa patients and non-mPCa patients or controls, respectively, in plasma and urinary cfDNA. (B) Determination of DMRs between PCa tissue and matched buffy coat samples revealed significant (adjusted p < 0.01), hyper- and hypomethylated regions in PCa. Identified 6015 hypermethylated regions (300 bp windows) with log2 fold change > 2 in PCa tissue compared to buffy coat were compared to TOP 100 hypermethylated DMRs (500 bp windows) obtained from an external data set (Börno et. al^17^), and 67 common regions (300 bp windows) were identified. Tissue-informed methylation markers were applied to cfMeDIP-seq data from LBx samples. (C) Left: Genomic annotation of 67 methylation marker regions; location within 3’ or 5’UTR, distal intergenic region, downstream region, exon or intron, promotor region. Right: Genomic annotation and assessment of CpG-associated landscapes (island, open sea, shelf, shore) within the 67 methylation marker regions. (D) Methylation scores (median of beta-values in 67 methylation marker regions) in plasma cfDNA (left) or urinary cfDNA (right) from cancer-free controls, lPCa patients, and aPCa patients. Each dot represents one sample. Box plot center lines indicate the median, and boxes illustrate the interquartile range with Tukey whiskers. The three cohorts were compared using Kruskal-Wallis testing, followed by Dunn’s post hoc test. Only significant differences are shown. adj. p = adjusted p value, aPCa = advanced prostate cancer, bp = base pair, cfDNA = cell-free DNA, DMR = differentially methylated region, LBx = Liquid Biopsy, lPCa = localized prostate cancer, M0 / M1 = absence/presence of distant metastases, mPCa = metastatic prostate cancer, n = number, PCa = prostate cancer, PTPRN2 = protein tyrosine phosphatase receptor type N2, SOX2-OT = SOX2 overlapping transcript, UTR = untranslated region, ZBTB46 = zinc finger and BTB domain containing 46

### PCa tissue methylation markers were elevated in LBx from PCa patients

Genome-wide differential methylation analysis was performed in PCa tissue and matched buffy coats to identify PCa-specific methylation patterns. Overall, 32,146 significant DMRs were identified, including 6015 hypermethylated (logFC > 2) and 1364 hypomethylated (logFC < -2) regions (**Fig. 1B**; *Supplementary Figure S2*). Most were located in introns and distal intergenic regions. Promoters accounted for 13% of hypermethylated and 6% of hypomethylated DMRs.

The 6015 hypermethylated DMRs were compared with a published MEDIP-seq dataset^17^, reporting 100 top-ranked DMRs (only hypermethylated) between 51 primary PCa and 53 normal prostate tissue samples. Sixty-seven shared regions were identified (**Fig. 1B**), predominantly located in introns (33%) and promoters (29%), with methylated CpGs mainly occurring inside CpG islands (**Fig. 1C**). Three intronic regions were associated with *PTPRN2*. Further validation using an external cfMeDIP-seq dataset of 133 plasma samples^20^ revealed that metastatic castration-resistant PCa (n = 103) clustered separately and demonstrated higher heterogeneity than localized PCa (lPCa, n = 30) based on methylation levels in these 67 regions, confirming their relevance as PCa biomarkers, particularly in advanced PCa (aPCa; *Supplementary Figure S3+S4*).

The 67 hypermethylated regions were analyzed in our cfMeDIP-seq data and a synoptic methylation score was calculated, showing an increasing trend from controls to lPCa to aPCa in both plasma and urine (**Fig. 1D**). Advanced PCa patients harbored significantly higher scores in urinary cfDNA; differences in plasma were not significant. The methylation score of 12 plasma (7x lPCa, 5x aPCa) and 12 urine samples (4x lPCa, 8x aPCa) exceeded the ctDNA detectability threshold. Five patients had elevated scores in matched plasma and urine, with higher levels in urine.

### Complementary genomic profiling in plasma and urine improved ctDNA detection

Genome-wide copy number variations (CNVs) were profiled in plasma and urinary cfDNA via lcWGS, assessing ctDNA presence and estimating tumorfractions.

CtDNA was mainly detected in either plasma or urine, indicating complementary CNV profiles (two examples in **Fig. 2A,B**). Five plasma samples (3x lPCa, 2x aPCa) and eight urine samples (5x lPCa, 3x aPCa) harbored ctDNA (*Supplementary Figure S5*).

**Fig. 2:**
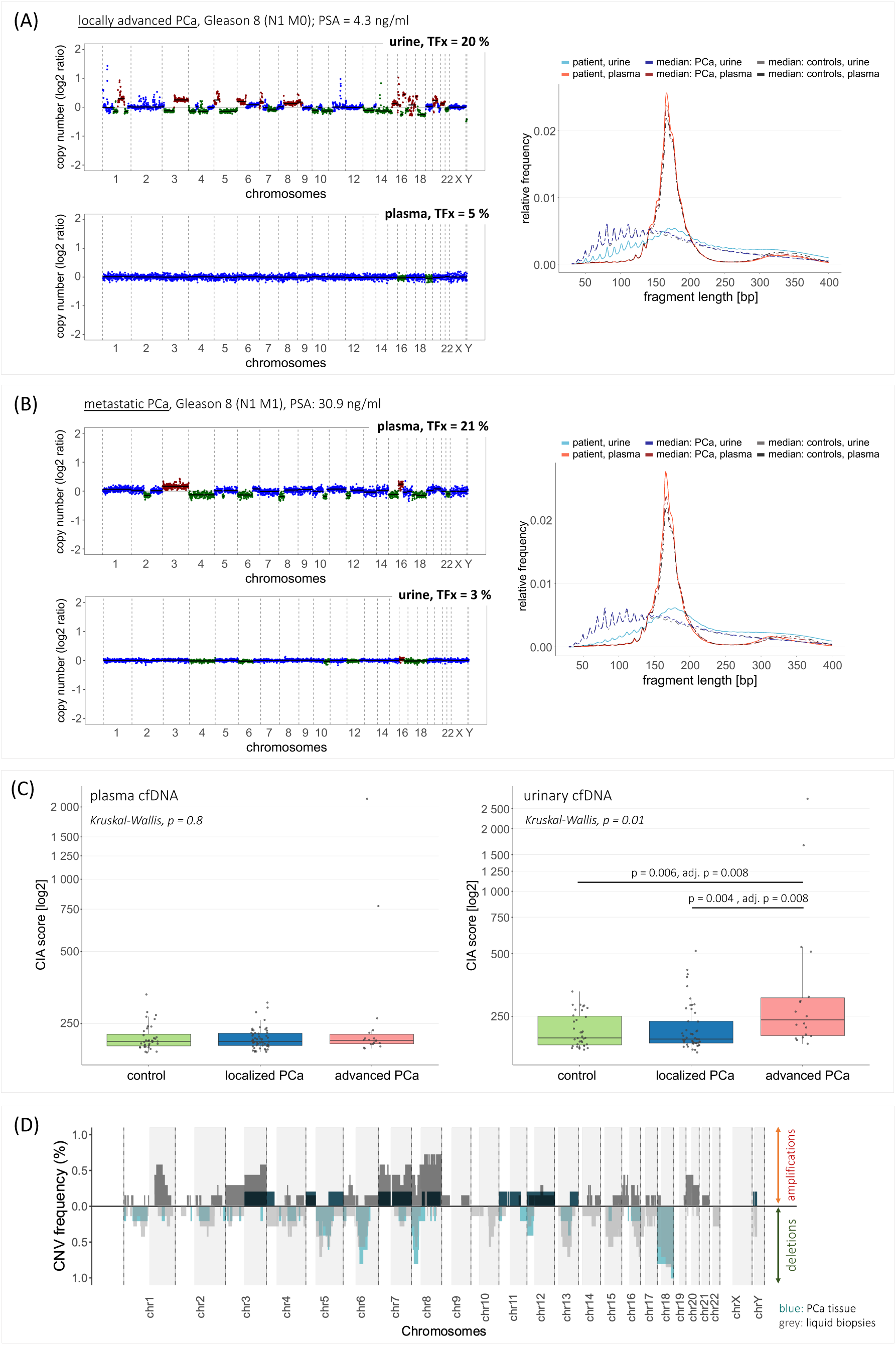
Genomic analysis in plasma and urinary cfDNA. (A) Complementary CNV profiles of plasma (left, bottom) and urine (left, top) samples from one patient with locally advanced PCa. Distinct CNVs were observed in the urine sample, while the plasma sample showed only few alterations. Right: Plasma and urinary cfDNA fragmentation profiles of the patient’s samples differed from the median profiles of all tumor and control samples. (B) Complementary CNV profiles of plasma (left, top) and urine (left, bottom) samples from one patient with mPCa. Distinct CNVs were observed in the plasma sample, while the urine sample showed only few alterations. Right: Plasma and urinary cfDNA fragmentation profiles of the patient’s samples differed from the median profiles of all tumor and control samples. (C) CIA scores in plasma cfDNA (left) or urinary cfDNA (right) from cancer-free controls, lPCa patients, and aPCa patients. Each dot represents one sample. Box plot center lines indicate the median, and boxes illustrate the interquartile range with Tukey whiskers. The three cohorts were compared using Kruskal-Wallis testing, followed by Dunn’s post hoc test. Only significant differences are shown. (D) Summary of recurrent amplifications and deletions in LBx samples (gray color) and eight PCa tissue samples (blue/turquoise color). Only LBx and PCa tissue samples with detectable CNVs and an estimated TFx > 10% were considered (PCa tissue: n = 5, LBx: n = 9). The y-axis indicates the frequency of a detected copy number state at the chromosomal coordinate specified on the x-axis across the samples. Areas shaded in gray represent the q-arm of the respective chromosome. chr = chromosome, CIA = chromosomal instability analysis, CNV = copy number variation, N0/N1 = absence/presence of lymph node metastases, TFx = tumorfraction

In-silico size selection (90–150 bp) aimed to enrich for ctDNA, as previously shown^12,21^, increasing the number of ctDNA-positive plasma samples to nine (5x lPCa, 4x aPCa). In urine, no optimal size selection was identified, due to heterogeneous cfDNA fragmentation (*Supplementary Figure S5*).

The CIA score served as additional genomic biomarker, targeting highly deviating regions to reduce background noise. Tumor samples had higher median CIA scores than controls in both fluids, with significant differences seen in urine between aPCa and controls or lPCa, respectively (both adjusted p = 0.008; **Fig. 2C**). Five plasma (3x lPCa, 2x aPCa) and 14 urine samples (7x lPCa, 7x aPCa), including one matched plasma-urine pair, exceeded the detectability threshold, indicating ctDNA presence.

LBx samples with TFx >10% harbored recurrent genomic alterations, consistent with our findings in eight PCa tissue samples (*Supplementary Figure S6*) and known alterations in primary PCa^2,22^. Deletions in 5q, 6q, 8p, 13q, and 18 were present in >40% of both tissue and LBx samples; gains in one tissue sample (3q, 5p, 7, 8q) were found in up to 60% of positive LBx (**Fig. 2D**).

### Plasma and urinary cfDNA fragmentation differed between PCa patients and controls

We analyzed global cfDNA fragmentation profiles and various fragment length ranges (*Supplementary Figures S7+S8*) to identify tumor-specific patterns and infer ctDNA presence. Plasma and urinary cfDNA fragmentation showed distinct characteristics, and differences between PCa patients and controls (**Fig. 3A,B)**. Plasma cfDNA fragmentation profiles exhibited a prominent 167 bp peak, representing nucleosomal DNA wrapping (plus linker DNA), and additional peaks (**Fig. 3A**; *Supplementary Figure S9*). Urinary cfDNA displayed a broader peak spanning 30–400 bp, though some samples contained an additional peak at ∼167 bp (**Fig. 3B**; *Supplementary Figure S10*). Both biofluids exhibited 10bp-oscillation patterns in fragments <150 bp (**Fig. 3A,B)**. In urine, this pattern was more pronounced and extended to longer fragments (150–300 bp; *Supplementary Figure S10*). Kolmogorov-Smirnov testing revealed significant differences in plasma cfDNA fragmentation between PCa and controls (p < 0.01), but not in urine due to higher inter-sample variability (*Supplementary Figures S9+S10*). The most tumor-informative plasma cfDNA fragmentation feature was the 10bp-oscillation pattern, with scores decreasing from controls to lPCa to aPCa (**Fig. 3C)**. Twelve tumor samples harbored 10bp-oscillation scores below the threshold (5^th^ percentile of controls), indicating ctDNA presence. In urine, PCa patients showed increased proportions of fragments with 163– 169 bp length (**Fig. 3C**), especially for aPCa compared to controls (adjusted p = 0.036), with one tumor sample exceeding the ctDNA detectability threshold.

**Fig. 3:**
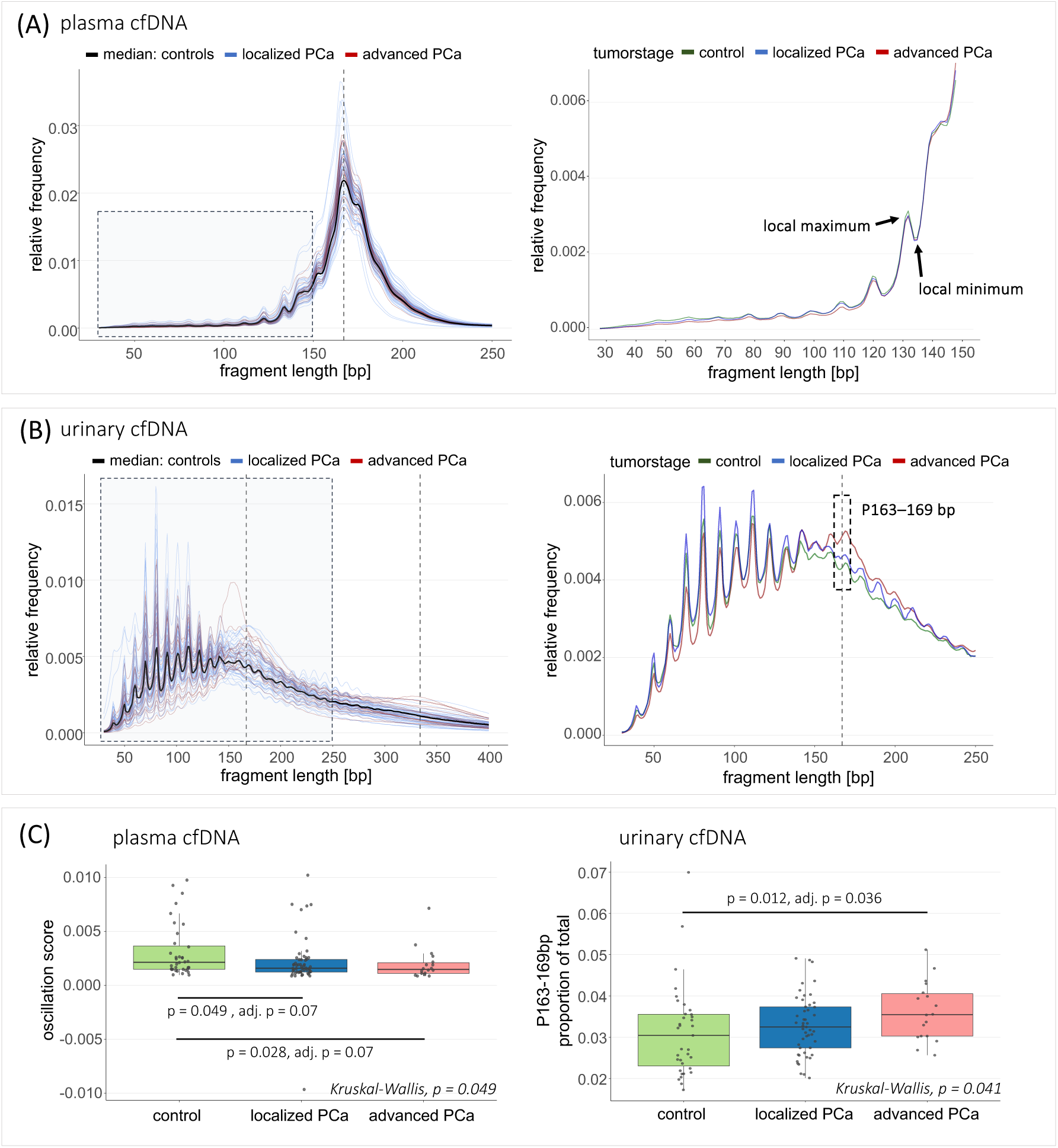
Fragmentation analysis of plasma and urinary cfDNA. (A) Left: Single plasma cfDNA fragmentation profiles of lPCa and aPCa patients, along with the median fragmentation profile of all cancer-free controls. Right: Plasma cfDNA fragmentation profiles (30–150 bp fragment length) exhibiting a 10bp-oscillation pattern with local maxima and minima at 10 bp intervals. Median fragmentation profiles are shown for all samples from lPCa patients, aPCa patients and cancer-free controls. (B) Left: Single urinary cfDNA fragmentation profiles of lPCa and aPCa patients, along with the median fragmentation profile of all cancer-free controls. Right: Urinary cfDNA fragmentation profiles, represented as median profiles of all samples from lPCa patients, aPCa patients and cancer-free controls. Fragment length range 163–169 bp is highlighted. (A+B) Y-axis: Relative frequencies of cfDNA fragments with specific length (bp) compared to all fragments (30–700 bp length). Vertical dotted grey line(s) indicate 167 bp and its multiple, 334 bp (2x167 bp). (C) Left: 10bp-oscillation scores (calculated based on the deviation between the sum of the height of all local maxima and the sum of the depth of all local minima) in plasma cfDNA from cancer-free controls, lPCa patients, and aPCa patients. Right: P163–169 bp values in urinary cfDNA from cancer-free controls, lPCa patients, and aPCa patients. Left, right: Box plot center lines indicate the median, and boxes illustrate the interquartile range with Tukey whiskers. Each dot represents one sample. The three cohorts were compared using Kruskal-Wallis testing, followed by Dunn’s post hoc test. Only significant differences are shown. P163–169 bp = proportion of fragments with length 163-169 bp in relation to all fragments

### Multimodal analyses enhanced ctDNA detection in PCa

Single-parameter analyses yielded low to moderate ctDNA detection rates, with plasma cfDNA fragmentation performing best in lPCa, followed by the CIA score in urine and the methylation score in plasma; other parameters showed detection rates ≤10% (**Fig. 4A**). In aPCa, detection rates increased across all features, with the methylation score and the CIA score in urine showing the best performance, followed by the methylation score, TFx, and cfDNA fragmentation in plasma (**Fig. 4A**). Complementary plasma-urine analysis improved ctDNA detection, yielding the highest detection rate based on CIA score and methylation score for lPCa and aPCa.

**Fig. 4:**
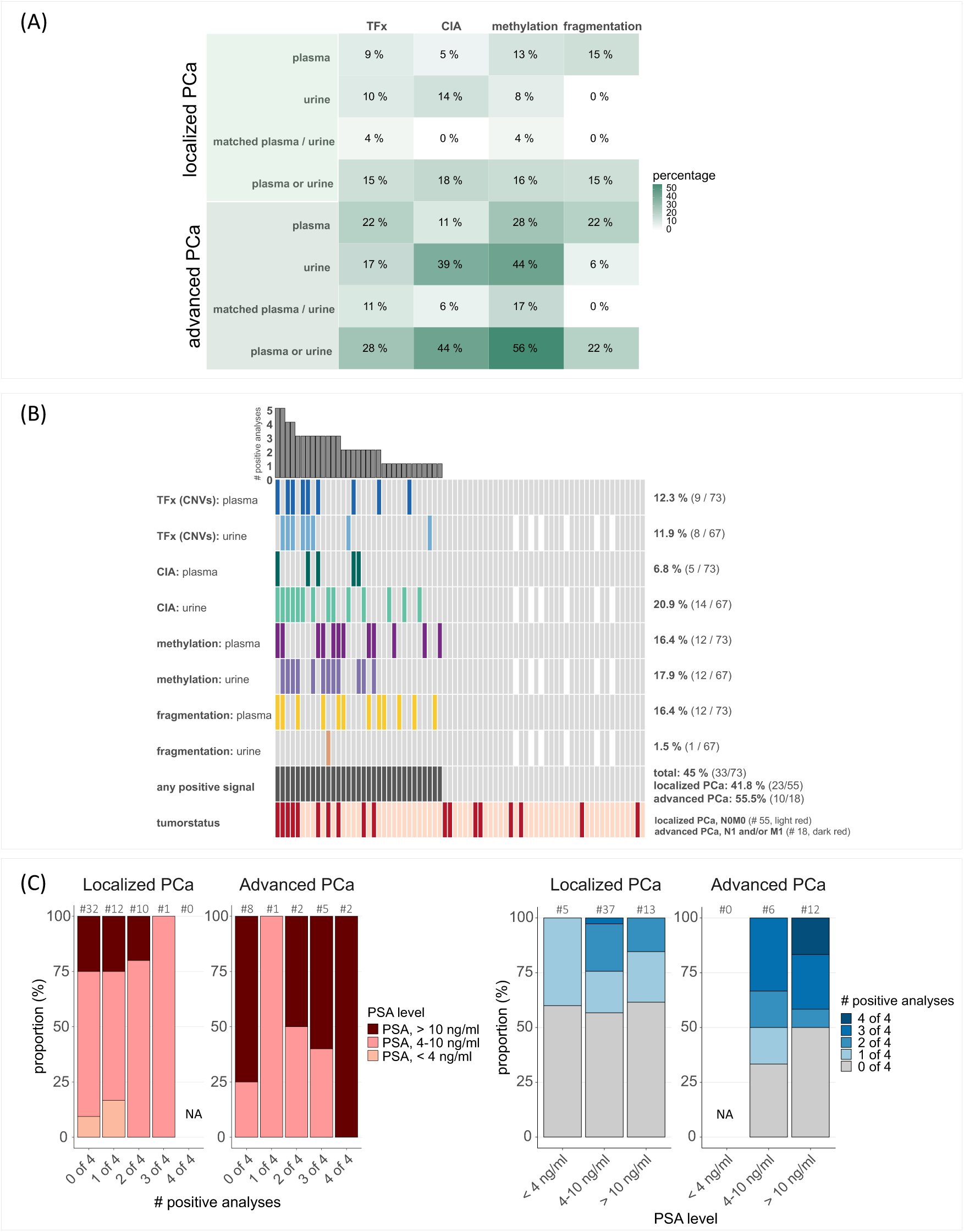
PCa detection and risk stratification based on multimodal LBx analyses. (A) Detection rates of tumor signals (ctDNA) in plasma and urine samples from lPCa (top) and aPCa (bottom) patients based on the four (epi)genomic analyses: 1) estimated TFx based on the CNV analysis with ichorCNA (plasma: analysis with in-silico size selection for 90–150bp fragments, urine: analysis without size selection), 2) CIA score, 3) methylation score, 4) cfDNA fragmentation (10bp-oscillation score in plasma cfDNA, P163–169 bp in urinary cfDNA). Detection rates (percentages, %) are shown separately for plasma and urine, in matched plasma and urine, and based on the complementary analysis combining results in both plasma and urine. Overall, 68 PCa patients harbored matched plasma and urine samples; 4 lPCa patients with missing urine samples. (B) Oncoprint with results from genomic (estimated TFx based on CNVs, CIA score) and epigenomic (cfDNA fragmentation features, methylation score) analyses in plasma and urinary cfDNA. Patients with detectable ctDNA in the respective analyses are indicated with colored tiles. White tiles represent non-available urine samples (n = 4). The bar plot on top depicts the number of positive analyses per patient. The second-to-last row represents the number of PCa patients with positive signal in at least one analysis (“any positive signal”; colored tiles). (C) Association between the number of positive LBx analyses and PSA levels for lPCa and aPCa patients. Left: Distribution of patients based on the number of positive tumor signals detected (0, 1, 2, 3, or 4 out of 4 analyses) in relation to PSA levels (< 4 ng/mL, 4–10 ng/mL, and > 10 ng/mL). Localized and aPCa are shown separately. Total numbers are depicted above each bar. Tumor signal positivity is determined using complementary analysis in plasma and urine across the four (epi)genomic assessments: TFx, CIA score, cfDNA fragmentation features, and methylation score. Right: Alternative representation of the data, displaying the number of patients within each PSA category (< 4 ng/mL, 4–10 ng/mL, and > 10 ng/mL) who show detectable ctDNA in 0,1,2,3 or 4 out of 4 analyses. Localized and aPCa are shown separately. Total numbers are depicted above each bar. ctDNA = circulating tumor-derived DNA, # = number of, PSA = prostate-specific antigen

Beyond complementarity of the two biofluids, combining genomic and epigenomic markers further enhanced ctDNA detection (**Fig. 4B**). In lPCa, ctDNA was detectable in 20% of plasma and 14% of urine samples based on one (epi)genomic feature, one urine sample was positive for three features, none for two or all four. In aPCa, 22% plasma samples were positive for one feature, 11% for two, and 6% for three and four features, respectively; 17% of urine samples were positive for one or three features, respectively. Considering all (epi)genomic cfDNA features in plasma and urine, ctDNA was detectable in 45% (33/73) of all PCa patients based on at least one positive result, including detection rates of 41.8% in lPCa and 55.5% in aPCa (**Fig. 4B**).

Furthermore, ctDNA was present in 43% of lPCa and 67% of aPCa cases with prostate-specific antigen (PSA) levels <10 ng/ml (**Fig. 4C**; *Supplementary Tables S3+S4*). Higher PSA levels correlated with an increasing number of positive analyses in both groups, especially in aPCa.

## 4 Discussion

This study evaluated genomic and epigenomic features in plasma and urinary cfDNA for ctDNA detection and risk stratification in newly diagnosed PCa. Our findings suggest that a multimodal approach provides a more comprehensive tumor characterization than single-parameter analyses. Complementary assessment of plasma and urine further enhanced sensitivity and achieved a ctDNA detection rate of 45%, underscoring the value of multi-source strategies. Epigenomic analyses revealed the best performance, with the 10bp-oscillation score in plasma cfDNA excelling in lPCa, and the methylation score in both fluids in aPCa. Genome-wide cfDNA methylation profiling identified hypermethylation in regions associated with *SOX2-OT*, *ZBTB46*, and *PTPRN2*, distinguishing mPCa from non-mPCa and controls. These genes are linked to tumorigenesis, androgen signaling, and metastasis^23–25^, reinforcing the value of methylation analysis for PCa classification and risk stratification. Genomic analyses contributed complementary information in plasma and urine for lPCa and aPCa, with increasing genomic instability observed with aPCa.

Multimodal LBx has potential to complement current diagnostics. Our findings indicated that higher PSA levels correlated with increased ctDNA detection, particularly in aPCa. For PSA >10 ng/ml, positive LBx findings could help distinguishing aggressive lPCa from aPCa, guiding risk-stratified treatment decisions toward surgery, multimodal therapy, or systemic treatment. CtDNA was also detectable in cases with PSA <10 ng/ml, emphasizing its promising diagnostic value for patients with inconclusive PSA values, refining risk stratification at multiple points. CfDNA analyses may support decision-making in cases of ambiguous imaging findings and guide further diagnostics, potentially reducing unnecessary procedures.

Our ctDNA detection rates aligned with prior studies, confirming higher sensitivities in mPCa^26^, while highlighting challenges in lPCa^7,8^. We demonstrate that multimodal analyses in two biofluids improved detection rates—a comprehensive approach not previously reported in newly diagnosed PCa. While complementary plasma-urine analyses are rare in PCa^27,28^, multimodal strategies have reinforced their potential for improved diagnostic accuracy in other malignancies^21,29–31^. Our analysis included (epi)genomic features previously shown to support tumor characterization. Mouliere et al. observed that cfDNA fragmentation patterns, including the 10bp-oscillation score, differed between tumor and control samples^12,32^. CNV analyses in plasma cfDNA using lcWGS were shown to detect ctDNA, predominantly in mPCa^8,33^. Methylation analyses showed promise for sensitive ctDNA detection through targeted approaches in plasma and urine from PCa patients^34,35^, as well as genome-wide profiling coupled with machine-learning classifiers which distinguished mPCa from lPCa or controls^20,36^.

Future research benefits from technological and bioinformatic advances to improve diagnostic sensitivity, particularly in early-stage PCa. Integrating multiple cfDNA parameters, as demonstrated in this study, or combining them with further data modalities (e.g., imaging) represent promising strategies for improved risk stratification, guiding clinical decision-making. CfDNA monitoring could refine therapeutic decisions at initial diagnosis and provide insights into treatment response and tumor progression for patients under systemic therapy. Larger prospective studies are essential to validate these findings and to assess their clinical applicability^37,38^.

Some limitations need to be addressed. The small sample size limited statistical power and generalizability of our findings. Although our approach improved ctDNA detection, sensitivities remained low, particularly for lPCa. Moreover, cfMeDIP-seq harbors limitations, including antibody-dependent enrichment biases, and underrepresentation of hypomethylation^14^.

## 5 Conclusion

Integrating multiple (epi)genomic cfDNA biomarkers from two LBx sources improved detection performance and risk stratification for PCa. Our findings supported the potential of multimodal LBx to complement PSA testing, particularly in cases with inconclusive PSA levels. The moderate costs and rapid turnaround time of cfDNA-based assays support their feasibility for clinical implementation. With ongoing technological progress, multimodal LBx approaches offer potential to improve clinical decision-making and contribute to future advancements in precision oncology, with possible translation to other urological malignancies.

## Data Availability

All data produced in the present study are available upon reasonable request to the authors.

## DECLARATIONS

### Conflicting interests

None.

### Funding

This work was realized through support by the German Federal Ministry for Economic Affairs and Climate Action (funding # 01MT21004A) and the Dieter Morszeck Foundation. The funders had no role in the design of the study; in the collection, analyses, or interpretation of data; in the writing of the manuscript, or in the decision to publish the results.

### Ethical approval

The study was approved by the ethical committee of the University of Heidelberg (Approval No. S-130/2021) and was performed in accordance with the Declaration of Helsinki.

### Contributorship

Conceptualization: A.L.R., M.G., H.S., F.J.; Data curation: A.L.R., S.E.; Formal analysis: A.L.R., S.E., F.J., O.L.; Funding acquisition: M.G.; Investigation: H.S., M.G., A.L.R.; Methodology: H.S., M.G., A.L.R., S.E., F.J.; Project Administration: M.G.; Resources: M.G., A.L.R.; Software: A.L.R., O.L., J.F.; Statistical analysis: A.L.R., S.E., F.J., O.L., D.H., O.S.; Supervision: M.G., H.S., D.H., O.S., S.D.; Visualization: A.L.R., F.J.; Writing - original draft: A.L.R.; Writing - review & editing: M.G., H.S., F.J.

All authors critically reviewed and approved the final version of the manuscript.

## Acknowledgements

The authors would like to thank all members of the Cancer Genome Research Group at German Cancer Research Center for their help and constructive discussion, in particular Arlou Angeles, Astrid Laut, Kate Glennon, Isabell Berneburg, Simon Ogrodnik and Sabrina Gerhardt. The authors would also like to thank the German Cancer Research Center core facilities for their support, in particular the NGS Core Facility for sequencing analyses, as well as the DKFZ Omics IT and Data Management Core Facility for data management and processing. Furthermore, we would like to thank the Tissue Bank of the National Center for Tumor Diseases (NCT) Heidelberg for their support in providing the tissue samples. We sincerely acknowledge our patients and their caregivers, as well as the help and support of the (medical) staff at the Urology Clinic of Heidelberg University Hospital.

## Data availability statement

The data sets generated and analyzed during the current study are available from the corresponding author on reasonable request.

## Declaration of generative AI and AI-assisted technologies in the writing process

During the preparation of this work, the authors used ChatGPT-4o in order to improve language and readability. After using this tool, the authors reviewed and edited the content as needed, and take full responsibility for the content of the publication.

## Notes

### Competing Interest Statement

The authors have declared no competing interest.

### Author Declarations

Ethics committee of the University of Heidelberg gave ethical approval for this work (Approval No. S-130/2021).

## References

1. Haffner MC, Zwart W, Roudier MP, et al. Genomic and phenotypic heterogeneity in prostate cancer. Nat Rev Urol. 2021;18(2):79–92. doi:10.1038/s41585-020-00400-w

2. Abeshouse A, Ahn J, Akbani R, et al. The Molecular Taxonomy of Primary Prostate Cancer. Cell. 2015;163(4):1011–1025. doi: 10.1016/j.cell.2015.10.025

3. Ali A, Du Feu A, Oliveira P, Choudhury A, Bristow RG, Baena E. Prostate zones and cancer: lost in transition? Nat Rev Urol. 2022;19(2):101–115. doi:10.1038/s41585-021-00524-7

4. De Mattos-Arruda L, Weigelt B, Cortes J, et al. Capturing intra-tumor genetic heterogeneity by de novo mutation profiling of circulating cell-free tumor DNA: a proof-of-principle. Ann Oncol. 2014;25(9):1729–1735. doi:10.1093/annonc/mdu239

5. Murtaza M, Dawson SJ, Pogrebniak K, et al. Multifocal clonal evolution characterized using circulating tumour DNA in a case of metastatic breast cancer. Nat Commun. 2015;6:8760. doi:10.1038/ncomms9760

6. Diaz LA, Jr., Bardelli A. Liquid biopsies: genotyping circulating tumor DNA. J Clin Oncol. 2014;32(6):579–86. doi:10.1200/JCO.2012.45.2011

7. Pope B, Park G, Lau E, et al. Ultrasensitive Detection of Circulating Tumour DNA enriches for Patients with a Greater Risk of Recurrence of Clinically Localised Prostate Cancer. Eur Urol. 2024;85(4):407–410. doi:h10.1016/j.eururo.2024.01.002

8. Hennigan ST, Trostel SY, Terrigino NT, et al. Low Abundance of Circulating Tumor DNA in Localized Prostate Cancer. JCO Precis Oncol. 2019;3. doi:10.1200/po.19.00176

9. Angeles AK, Janke F, Bauer S, Christopoulos P, Riediger AL, Sültmann H. Liquid Biopsies beyond Mutation Calling: Genomic and Epigenomic Features of Cell-Free DNA in Cancer. Cancers (Basel). 2021;13(22):5615. doi: 10.3390/cancers13225615

10. van der Pol Y, Mouliere F. Toward the Early Detection of Cancer by Decoding the Epigenetic and Environmental Fingerprints of Cell-Free DNA. Cancer Cell. 2019;36(4):350–368. doi:10.1016/j.ccell.2019.09.003

11. Janke F, Gasser M, Angeles AK, et al. Low-coverage whole genome sequencing of cell-free DNA to predict and track immunotherapy response in advanced non-small cell lung cancer. J Exp Clin Cancer Res. 2025;44(1):87. doi:10.1186/s13046-025-03348-0

12. Mouliere F, Chandrananda D, Piskorz AM, et al. Enhanced detection of circulating tumor DNA by fragment size analysis. Sci Transl Med. 2018;10(466). doi:10.1126/scitranslmed.aat4921

13. Moldovan N, van der Pol Y, van den Ende T, et al. Multi-modal cell-free DNA genomic and fragmentomic patterns enhance cancer survival and recurrence analysis. Cell Rep Med. 2024;5(1):101349. doi:10.1016/j.xcrm.2023.101349

14. Shen SY, Burgener JM, Bratman SV, De Carvalho DD. Preparation of cfMeDIP-seq libraries for methylome profiling of plasma cell-free DNA. Nat Protoc. 2019;14(10):2749–2780. doi:10.1038/s41596-019-0202-2

15. Adalsteinsson VA, Ha G, Freeman SS, et al. Scalable whole-exome sequencing of cell-free DNA reveals high concordance with metastatic tumors. Nat Commun. 2017;8(1):1324. doi:10.1038/s41467-017-00965-y

16. Chemi F, Pearce SP, Clipson A, et al. cfDNA methylome profiling for detection and subtyping of small cell lung cancers. Nat Cancer. 2022;3(10):1260–1270. doi:10.1038/s43018-022-00415-9

17. Börno ST, Fischer A, Kerick M, et al. Genome-wide DNA methylation events in TMPRSS2-ERG fusion-negative prostate cancers implicate an EZH2-dependent mechanism with miR-26a hypermethylation. Cancer Discov. 2012;2(11):1024–35. doi:10.1158/2159-8290.CD-12-0041

18. R Foundation for Statistical Computing. R: A Language and Environment for Statistical Computing. 2020. https://www.R-project.org/

19. Wickham H. ggplot2: Elegant Graphics for Data Analysis. Springer-Verlag New York. 2016. https://ggplot2.tidyverse.org

20. Chen S, Petricca J, Ye W, et al. The cell-free DNA methylome captures distinctions between localized and metastatic prostate tumors. Nat Commun. 2022;13(1):6467. doi:10.1038/s41467-022-34012-2

21. Smith CG, Moser T, Mouliere F, et al. Comprehensive characterization of cell-free tumor DNA in plasma and urine of patients with renal tumors. Genome Med. 2020;12(1):23. doi:10.1186/s13073-020-00723-8

22. Taylor BS, Schultz N, Hieronymus H, et al. Integrative genomic profiling of human prostate cancer. Cancer Cell. 2010;18(1):11–22. doi:10.1016/j.ccr.2010.05.026

23. Kar S, Niharika, Roy A, Patra SK. Overexpression of SOX2 Gene by Histone Modifications: SOX2 Enhances Human Prostate and Breast Cancer Progression by Prevention of Apoptosis and Enhancing Cell Proliferation. Oncology. 2023;101(9):591–608. doi:10.1159/000531195

24. Gentilini D, Scala S, Gaudenzi G, et al. Epigenome-wide association study in hepatocellular carcinoma: Identification of stochastic epigenetic mutations through an innovative statistical approach. Oncotarget. 2017;8(26):41890–41902. doi:10.18632/oncotarget.17462

25. Chen WY, Tsai YC, Siu MK, et al. Inhibition of the androgen receptor induces a novel tumor promoter, ZBTB46, for prostate cancer metastasis. Oncogene. 2017;36(45):6213–6224. doi:10.1038/onc.2017.226

26. Wyatt AW, Annala M, Aggarwal R, et al. Concordance of Circulating Tumor DNA and Matched Metastatic Tissue Biopsy in Prostate Cancer. J Natl Cancer Inst. 2017;109(12)doi:10.1093/jnci/djx118

27. Silva R, Moran B, Russell NM, et al. Evaluating liquid biopsies for methylomic profiling of prostate cancer.Epigenetics.2020;15(6-7):715–727.doi:10.1080/15592294.2020.1712876

28. Chen G, Jia G, Chao F, et al. Urine- and Blood-Based Molecular Profiling of Human Prostate Cancer. Original Research. Front Oncol. 2022;12. doi:10.3389/fonc.2022.759791

29. Peneder P, Stütz AM, Surdez D, et al. Multimodal analysis of cell-free DNA whole-genome sequencing for pediatric cancers with low mutational burden. Nat Commun. 2021;12(1):3230. doi:10.1038/s41467-021-23445-w

30. Burgener JM, Zou J, Zhao Z, et al. Tumor-Naive Multimodal Profiling of Circulating Tumor DNA in Head and Neck Squamous Cell Carcinoma. Clin Cancer Res. 2021;27(15):4230–4244. doi:10.1158/1078-0432.CCR-21-0110

31. Cheng THT, Jiang P, Teoh JYC, et al. Noninvasive Detection of Bladder Cancer by Shallow-Depth Genome-Wide Bisulfite Sequencing of Urinary Cell-Free DNA for Methylation and Copy Number Profiling. Clin Chem. 2019;65(7):927–936. doi:10.1373/clinchem.2018.301341

32. Mouliere F, Smith CG, Heider K, et al. Fragmentation patterns and personalized sequencing of cell-free DNA in urine and plasma of glioma patients. EMBO Mol Med. 2021; 13(8):e12881. doi:10.15252/emmm.202012881

33. Heitzer E, Ulz P, Belic J, et al. Tumor-associated copy number changes in the circulation of patients with prostate cancer identified through whole-genome sequencing. Genome Med. 2013;5(4):30. doi:10.1186/gm434

34. Payne SR, Serth J, Schostak M, et al. DNA methylation biomarkers of prostate cancer: confirmation of candidates and evidence urine is the most sensitive body fluid for non-invasive detection. Prostate. 2009;69(12):1257–69. doi:10.1002/pros.20967

35. Klein EA, Richards D, Cohn A, et al. Clinical validation of a targeted methylation-based multi-cancer early detection test using an independent validation set. Ann Oncol. 2021;32(9):1167–1177. doi:10.1016/j.annonc.2021.05.806

36. Lleshi E, Milne-Clark T, Lee Yu H, et al. Prostate cancer detection through unbiased capture of methylated cell-free DNA. iScience.2024;27(7):110330. doi:10.1016/j.isci.2024.110330

37. Alix-Panabières C, Marchetti D, Lang JE. Liquid biopsy: from concept to clinical application. Sci Rep. 2023;13(1):21685. doi:10.1038/s41598-023-48501-x

38. Edsjö A, Holmquist L, Geoerger B, et al. Precision cancer medicine: Concepts, current practice, and future developments. J Intern Med. 2023;294(4):455–481. doi:10.1111/joim.13709

39. TNM classification of malignant tumours. 8th Edition ed. UICC. John Wiley & Sons; 2016.

40. D’Amico AV, Whittington R, Malkowicz SB, et al. Biochemical outcome after radical prostatectomy, external beam radiation therapy, or interstitial radiation therapy for clinically localized prostate cancer. Jama. 1998;280(11):969–74. doi:10.1001/jama.280.11.969

