## Supplementary Material for "Integrative genomic and epigenomic profiling in plasma and urinary cell-free DNA improves early risk stratification of newly diagnosed prostate cancer"

**Supplementary Text**

Supplementary Patients and Methods

Supplementary Note

**Supplementary Figures and Tables**

**References**

### Supplementary Patients and Methods

Patients and cancer-free controls

This study included 73 prostate cancer (PCa) patients and 36 cancer-free controls recruited at the Urology Clinic at Heidelberg University Hospital between June 2021 and November 2022, with ethics approval (ethic committee of the Medical Faculty of Heidelberg University; S-130/2021) and informed consent obtained. PCa patients were enrolled at the time of initial diagnosis and underwent prostate biopsy and/or radical prostatectomy at the University Hospital Heidelberg. In case of 47 men, blood and urine samples were collected before prostate tissue biopsy, which subsequently confirmed PCa diagnosis. One patient underwent two biopsies within a year, with liquid biopsy (LBx) samples taken at both time points, revealing a re-classification from Gleason 6 to Gleason 7b. Additionally, 25 PCa patients diagnosed through internal or external biopsy provided LBx samples few weeks after diagnosis and prior to radical prostatectomy. The cancer-free control cohort included 16 men with PSA > 2 ng/ml who had a prostate biopsy at Heidelberg University Hospital with no evidence of malignancy, alongside 14 men with PSA < 2 ng/ml who underwent surgeries or examinations due to benign urological conditions at Heidelberg University Hospital. Comprehensive clinical data were available for all men and PSA levels were obtained from internal measurements or external documentation. Multiparametric magnetic resonance imaging (MRI) data were available for most PCa patients and biopsy-negative controls. Multiparametric MRI was performed at the Radiological department of the German Cancer Research Center (DKFZ) Heidelberg or external centers. Multiparametric prostate MRI was assessed following the Prostate Imaging-Reporting and Data System (PI-RADS) recommendations[1]. Externally conducted MRI scans were again reviewed internally.

Prostate biopsies at the Urology Clinic Heidelberg were performed as transperineal fusion-targeted biopsies of PI-RADS 3–5 lesions, with elastic software registration using UroNav (Philips Invivo, Gainesville, FL, USA), combined with volume-adjusted systematic saturation biopsies, as previously described[2,3]. Histopathological assessments were performed at the Institute of Pathology of Heidelberg University Hospital, adhering to International Society of Urological Pathology standards[4].

Sample collection, processing and DNA extraction

*LBx samples*

Blood samples were drawn via cubital puncture between 8 and 11 AM from fasting patients (nine individuals were non-fasting). Two additional 9 mL EDTA blood tubes (Sarstedt S-Monovette K3) were collected for biobanking, alongside routine laboratory assessments. Patients provided 30–50 mL midstream urine in sterile 200 mL beakers. One part of the volume was used for routine tests (urine status and -culture) and the remaining volume was stored in 1–2 x 15 mL falcon tubes, containing 200 µL of 0.5 M EDTA (pH 8.0) for stabilization. Blood and urine samples were kept at 4°C and further processed within 4–6 hours. Blood samples were centrifuged at 1,600 × g for 10 minutes at 4°C. Plasma aliquots (2 × 3.2–4.8 mL) were again centrifuged at 14,000 × g for 10 minutes at 4°C to remove cell debris. Buffy coats were equally obtained and stored in 2 mL tubes. Urine samples underwent an initial centrifugation at 750 × g for 10 minutes at 4°C, followed by a second spin at 2,600 × g to further clear cell debris. All centrifugation steps were conducted without brake. All samples were stored at −80°C until further use.

Cell-free DNA (cfDNA) was extracted from 1–5.5 ml of plasma and 9.5–19 ml of urine using the QIAamp MinElute ccfDNA Kit (Qiagen), following the manufacturer’s protocol with minor modifications. Frozen samples were thawed on ice and centrifuged at 1,500 × g for 2 minutes at room temperature to remove residual cell debris. Reagents were prepared according to the manufacturer´s instructions, with adjustments based on sample volume. Briefly, proteinase K, magnetic bead suspension, and bead binding buffer were added in volume-adjusted amounts, followed by 10-minute incubation with end-over-end rotation. Bead-bound cfDNA was precipitated using a magnetic rack and eluted in 200 µL of bead elution buffer (350 µL for urine), then incubated at 300 rpm for 5 minutes to release cfDNA. After capturing the empty beads, the cfDNA-containing supernatant was combined with 300 µL Buffer ACB (400 µL for urine) and loaded onto QIAamp UCP MinElute columns. Samples were centrifuged at 6,000 × g for 1 minute, followed by a single wash with Buffer ACW2 and an additional 3-minute centrifugation at 20,000 × g to dry the silica membrane. Final elution was performed in 30–50 µL of nuclease-free water, and cfDNA was stored at −20°C until further analysis. CfDNA concentrations were measured using the Qubit 2.0 Fluorometer with the Qubit dsDNA High-Sensitivity (HS) Assay Kit, following the manufacturer's protocol for sample preparation and instrument loading. Fragment size distribution and DNA integrity were analyzed using the Fragment Analyzer 5200 (Agilent) with the HS NGS Fragment Kit (1–6000 bp). Measurements followed the manufacturer's guidelines, and fragment length distributions were assessed using the ProSize data analysis software v4.0.1.4 in NGS analysis mode.

Genomic DNA (gDNA) was extracted from eight buffy coat samples using the QIAamp DNA Mini and Blood Mini Kit (Qiagen), following the manufacturer's protocol. DNA quantification was performed with the NanoDrop 1000 Spectrophotometer, and fragment length distribution was assessed using the Fragment Analyzer 5200 (Agilent) with the NGS Fragment Kit (1–6000 bp). Extracted gDNA from buffy coat samples (1 µg of gDNA input) was sheared using the Covaris M220 Focused Ultrasonicator to generate uniform DNA fragments for NGS library preparation. Shearing followed the manufacturer's protocol with the following settings: PIP: 50 W, Duty Factor: 20%, Cycles per Burst: 200, Time: 220 sec, producing fragments of ~150–220 bp. Sheared gDNA was quantified with the Qubit 2.0 Fluorometer (Qubit dsDNA HS Assay Kit) and the fragment size distribution was re-assessed with the Fragment Analyzer 5200 (HS NGS Fragment Kit, 1–6000 bp), as previously described.

*Fresh-frozen PCa tissue samples*

PCa tissue samples were obtained from eight patients (matching LBx samples were also analyzed) undergoing radical prostatectomy at Heidelberg University Hospital. Extracted intraoperatively, the tissue samples (~80 mg) were cryopreserved in liquid nitrogen and stored at −80°C until further processing in the Tissue Bank of the National Center for Tumor Diseases (NCT) Heidelberg, in accordance with the regulations of the tissue bank and the approval of the ethics committee of Heidelberg University. Histopathological assessment was conducted at the Institute of Pathology of Heidelberg University Hospital, where tissue quality and tumor cell content were evaluated. The tissue samples contained 40–90 % tumor cells.

Genomic DNA was extracted from eight fresh-frozen PCa tissue samples using the AllPrep DNA/RNA/Protein Mini Kit (Qiagen) for simultaneous DNA, RNA, and protein purification. The procedure followed the manufacturer's instructions. Extracted gDNA samples were quantified using the NanoDrop 1000 Spectrophotometer and analyzed for fragment length distribution with the Fragment Analyzer 5200, applying the NGS Fragment Kit (1–6000 bp), following the manufacturers' protocols. Shearing of extracted gDNA from fresh-frozen PCa tissue samples was performed with use of the Covaris M220 Focused ultrasonicator, as already described.

Low-coverage whole-genome sequencing (lcWGS) and (cell-free) methylated DNA immunoprecipitation sequencing ((cf)MeDIP-seq)

*LBx samples (cfDNA)*

Genome-wide methylation profiling of cfDNA from 109 plasma and 102 urine samples was performed using cfMeDIP-seq, following the protocol of Shen et al. [5] with adaptions for local implementation. The workflow included library preparation, methylation enrichment, and the generation of non-enriched libraries for lcWGS. Library preparation involved end-repair, A-tailing, and adapter ligation using the KAPA Hyper Prep Kit (Roche), according to the manufacturer´s protocol. For cfDNA samples, 7 ng input was generally used. In cases with lower total cfDNA quantities available, inputs ≥2.49 ng yielded adequate libraries. Adapter ligation was performed overnight with NEBNext UDI-UMI adaptors (New England Biolabs). Adapter-ligated DNA underwent bead purification and double-sided size selection (>50–100 bp, <800 bp) with AMPure XP beads. To adapt the methylation enrichment protocol to low cfDNA input quantities[5], pre-synthesized filler DNA (λ DNA) of varying methylation levels was added to maintain a final quantity of 100 ng total DNA. Amount of filler DNA was adjusted according to the initial cfDNA input.

The MeDIP workflow included i) addition of methylated (5mC)- and unmethylated (5C)- spike-ins for methylation quality control (QC), ii) sample denaturation, iii) separation of input control (non-enriched sample) and immunoprecipitated (enriched) sample, and iv) methylation enrichment, using the MagMeDIP qPCR Kit (Diagenode), according to the manufacturer´s protocol. At the beginning of the protocol, 5mC- and 5C- spike-ins were added to each sample. Both 5mC-/5C-spike-ins and their respective quantitative polymerase chain reaction (qPCR) primers were supplied in the MagMeDIP qPCR kit. Before methylation enrichment, 20% of the sample was set aside as a non-enriched control. The remaining 80% underwent overnight incubation with a 5mC-specific antibody, followed by washing and DNA purification using the iPure Kit (Diagenode). Enriched and non-enriched DNA were purified in parallel, generating two libraries per sample. The non-enriched fraction (~2 ng cfDNA) served as a QC reference for methylation enrichment and was used for lcWGS. Final library amplification was performed using 12–13 PCR cycles for enriched libraries and 8–9 cycles for non-enriched libraries. Fragment size distributions of all libraries were analyzed with the Fragment Analyzer 5200 (NGS Fragment Kit, 1–6000 bp) and quantified using the Qubit 2.0 Fluorometer (dsDNA HS Assay Kit). Equimolar pooling (10 µM) was performed for multiplexed 100 bp paired-end sequencing on the Illumina NovaSeq 6000 platform. Sequencing, flow-cell loading, and cluster formation were conducted at the DKFZ Genomics and Proteomics Core Facility, following standard protocols. LcWGS libraries were sequenced to a median coverage of 2.76× (1.38–4.28×) and cfMeDIP-seq libraries had a median coverage in methylation-enriched regions of 2.73× (range 0.002–6.63×; **Supplementary Table S1**).

*Fresh-frozen tissue and matched buffy coat samples (gDNA)*

The same library preparation protocol was applied, with few modifications, to eight fresh-frozen tissue and matched buffy coat samples using sheared gDNA (100 ng) as input. Consequently, no filler DNA was added to the samples and the molarity of NEBNext UDI-UMI adaptors (New England Biolabs) for adapter ligation was adjusted to increased input amount. The non-enriched libraries, comprising 20% of the original volume, contained ~20 ng of sheared gDNA. Due to the higher initial gDNA input, enriched and non-enriched libraries were amplified with only 8 and 6 PCR cycles, respectively. Final MeDIP-seq and lcWGS libraries were sequenced alongside libraries from LBx samples. LcWGS libraries were sequenced to a median coverage of 2.32× (range 2.07–2.66×) and cfMeDIP-seq libraries had a median coverage in methylation-enriched regions of 2.25× (range 1.51–3.27×; **Supplementary Table S1**).

<u>Processing of raw sequencing data</u>

Raw sequencing data from (cf)MeDIP-seq and lcWGS libraries were provided as demultiplexed FASTQ files (R1, R2, UMI information) by the High Throughput Sequencing Unit of the DKFZ Genomic and Proteomics Core Facility. Data preprocessing and alignment involved adapter trimming, read alignment to the human reference genome (hg19), duplicate removal, and quality filtering. Data processing was performed using an in-house Nextflow pipeline[6]. Software dependencies were managed via Singularity containers[7], and workflow components were sourced from nf-core[8], with additional custom modules and sub-workflows.

The first processing step involved UMI extraction from the third FASTQ file using UMI-tools v1.1.2[9], appending the barcode to the read name. Remaining Illumina adapter sequences were removed with cutadapt v3.4[10], and trimmed reads were aligned to hg19 (UCSC genome browser) using Bowtie2 v2.4.4[11] with default parameters, restricting insert sizes to 30–700 bp. The resulting SAM files were converted to sorted, indexed BAM files via Samtools v1.15.1[12]. Quality filtering with Samtools v1.15.1[12] retained only properly paired reads which were mapped in proper pairs, excluding unmapped reads or reads with unmapped mate, with a mapping quality threshold (MAPQ > 10). Deduplication was performed with UMI-tools v.1.1.2[9] (network-based method; default), removing redundant reads while preserving one representative read per UMI group. Unmapped, unpaired, and chimeric reads were discarded. Final BAM files were sorted and indexed with Samtools v1.15.1[12]. QC was conducted at multiple stages. FastQC v0.11.9[13] assessed raw sequencing reads (FASTQ), aligned reads, and final processed data. Additionally, Samtools v1.15.1[12] (stats, flagstat, idxstat) provided summary statistics for aligned and filtered reads. All QC results were compiled using MultiQC v1.12[14]. Methylation enrichment efficacy was evaluated through bioinformatic analyses of processed (cf)MeDIP-seq data, including assessments of the saturation, CpG coverage, and (relative) methylation enrichment score using the R package MEDIPS v.1.46.0[15].

Genome-wide methylation profiling

Genome-wide methylation profiling and assessment of differentially methylated regions (DMRs) were performed using (cf)MeDIP-seq data from plasma or urine, as well as PCa tissue and matched buffy coat samples. Data processing and DMR identification were conducted with R package MESA v0.2.2[16], built on R package QSEA v1.16.0[17], with adaptations for compatibility with GRCh37/hg19.

In the first step, a QSEAset was created based on pre-processed BAM files, storing metadata, genome-wide coverage in 300 bp bins, copy number variation (CNV) data (log fold change (logFC) in 1 Mb windows), and sequencing parameters, including normalization factors. Read coverage and CpG density were computed per genomic window, while ENCODE-blacklisted regions[18], reads with mapping quality <10, and low-coverage windows (<10 reads across samples) were excluded. Library size normalization was performed using TMM (trimmed mean of M-values), and CNV correction was applied with CNV data extracted from paired non-enriched lcWGS data. Genome-wide, relative methylation signals were expressed as normalized reads per kilobase million (nrpkm), representing counts in 300 bp windows adjusted for CNVs, library size, region length, and zygosity. Absolute methylation levels for each window were estimated as β-values (scaled 0–1, where 0 is unmethylated and 1 is fully methylated), using the "blind calibration" method from the QSEA algorithm.

*Determination of DMRs in LBx samples*

DMRs were identified in plasma and urinary cfDNA by comparing cfMeDIP-seq data from all tumor patients, tumor sub-cohorts, and controls. Two QSEAsets (one for plasma and urine, respectively) were created, and subsets were extracted for specific comparisons. DMR analysis was performed using R package MESA v0.2.2[16].

Low-coverage 300 bp windows were excluded from the DMR analysis, in which no sample exceeded two NRPKMs. Absolute methylation levels (β-values) in the remaining windows were compared between cohorts using a generalized linear model (GLM) with a negative binomial distribution and a logarithmic link function. Significance was tested by comparing a reduced model (normal vs. tumor) to a Chi-squared distribution, with Benjamini-Hochberg correction ($q < 0.05$) for multiple testing. Methylation differences were expressed as logFC, where positive logFC indicated hypermethylation and negative logFC hypomethylation relative to the reference group (e.g., controls). DMRs were visualized with volcano plots, displaying logFC on the x-axis and -log10(adjusted p-value) on the y-axis. Significant, hypermethylated (logFC > 1, $q < 0.05$) and hypomethylated DMRs (logFC < -1, $q < 0.05$) were highlighted in colors. Hierarchical clustering of β-values from tumor and control samples was conducted with R package pheatmap v.1.0.12[19]. Furthermore, DMRs were annotated for genomic regions (e.g., promoters, exons, introns, UTRs) and CpG landscapes (CpG islands, shores, shelves, open sea) using an annotation function included in R package MESA v0.2.2[16] (based on R package ChIPseeker[20]). The distribution of annotated features was visualized with bar plots and pie charts in R package ggplot2 v3.4.2[21]. Overlapping DMRs between cohorts or between plasma and urine samples were identified with R package ChIPpeakAnno v.3.24.2[22].

*Genome-wide methylation profiling in PCa tissue samples and assessment of PCa-specific methylation markers in LBx samples*

Genome-wide methylation profiling was also conducted on eight PCa tissue samples using MeDIP-seq data. DMRs were identified in 300 bp windows by comparing PCa tissue with matched buffy coat samples as a normal reference. DMR analysis was performed with R package MESA v0.2.2[16], as already described. Significant DMRs were defined using $q < 0.01$, and filtered for $logFC > 2$ (hypermethylated) or $logFC < -2$ (hypomethylated) in PCa tissue. Results were visualized in volcano plots, and DMRs were annotated for genomic regions and CpG landscapes. To validate findings and identify PCa-specific methylation markers for LBx analysis, an external MeDIP-seq dataset from Börno et al. (51 primary PCa and 53 normal prostate tissue samples) was used[23]. The top 100 hypermethylated DMRs (500 bp windows), ranked by the lowest p-values, from the Supplementary Data (Table S3a) of the study[23] were overlapped with significant, hypermethylated DMRs ($logFC > 2$) from our PCa tissue analysis using R package ChIPpeakAnno v.3.24.2[22], and the overlap was visualized in a Venn diagram. The common 300 bp regions were selected as methylation markers for LBx analysis. For application on LBx data, β-values in the selected 300 bp-regions were extracted for plasma and urine samples from the own study cohort, and statistically compared between tumor and control samples. A methylation score was calculated for each plasma and urine sample as the median β-value across the selected regions, serving as a synoptic measure of the methylation status in PCa patients and controls.

Additionally, methylation markers were validated in an external cfMeDIP-seq dataset from Chen et al. [24], including 133 plasma samples from localized PCa (lPCa, n = 30) and metastatic castration-resistant PCa (mCRPC, n = 103). The dataset was obtained from the European Genome-Phenome Archive (EGA): EGAD00001007972, EGAD00001008711,

EGAD00001008712, EGAD00001008713, EGAD00001008737. Plasma samples from lPCa were sourced from the Canadian Prostate Cancer Genome Network (CPC-GENE) project[25], while mCRPC samples were obtained from three cohorts (Barrier, WCDT, and VPC), comprising patients who underwent androgen receptor inhibition therapy[26,27]. Additionally, a validation cohort with 72 plasma samples from the VPC cohort was collected (EGAD00001008737), but was ultimately excluded from our own comparative analysis. External FASTQ files were processed using our in-house Nextflow pipeline (previously described), excluding UMI-based deduplication (as UMI adaptors were not used in the external data set) and instead using Picard v.2.27.2[28] to mark and remove duplicates. A QSEAset was generated for cfMeDIP-seq data from lPCa and mCRPC patients using the R package MESA v0.2.2[16], applying the same parameters as for the own study's LBx samples. Since non-enriched WGS data was not available for CNV correction, CNVs were estimated from cfMeDIP-seq data. Hierarchical clustering of β-values in the selected regions based on our tissue analysis was performed using R package pheatmap v.1.0.12, and principal component analysis (PCA) was conducted using R package stats v.4.0.0[29]. PCA plots were generated with R packages plotly v.4.10.2[30] and ggfortify v.0.4.17[31], displaying the first two principal components with their explained variance.

<u>Genome-wide analysis of CNVs and chromosomal instability</u>

*LBx samples (cfDNA)*

Genome-wide CNV profiling of plasma and urinary cfDNA was performed using lcWGS data. The analysis was implemented as a Nextflow pipeline incorporating necessary processing steps and ichorCNA v.0.4.0[32] for the CNV analysis, with software dependencies managed via Singularity[7]. Several modules generated by the nf-core community[8] were integrated, with minor adaptations. In the first step, sequencing reads were filtered to remove ENCODE-blacklisted[18] genomic regions. Coverage was standardized by downsampling to 20 million paired reads using Samtools v1.15.1[12]. Genomes were segmented into 1000 kilobase (kb) bins using HMM Copy Utils[33,34], with read counts stored in WIG format for input into the ichorCNA algorithm. Normalization for GC content and mappability was applied using HMMcopy v1.32.0[34]. One Panel of Normals (PoN) was generated by combining all control samples as a copy-neutral reference for tumor samples, while individual PoNs were created for each control sample by excluding the analyzed control sample from the PoN for tumor samples. The ichorCNA workflow used a hidden Markov model (HMM) for CNV segmentation, subsequent prediction of large-scale CNVs, and estimation of the tumorfraction (TFx). Parameter settings of the ichorCNA algorithm were adjusted to account for the expected low tumor content in cfDNA. All chromosomes were analyzed, but only autosomes were used for parameter estimation. Since all patients and controls were male, gender was set to "male".

CNV analysis was conducted on all sequencing reads (30–700 bp), followed by an additional in-silico size selection. Due to the significant reduction in reads following size selection, downsampling was adjusted to 2 million paired reads. For plasma cfDNA, size selection was applied to the 90–150 bp fragment length range, previously shown to enhance ctDNA detection in CNV analysis[35,36]. For urinary cfDNA, different size ranges were tested to determine the most effective range for improving CNV detection, including 90–150 bp, 110–170 bp, 20–100 bp, 20–110 bp, 20–120 bp, 20–130 bp, 20–140 bp, 20–150 bp, 20–160 bp, 20–170 bp, 140–170 bp, 140–180 bp, 140–190 bp, 140–200 bp, and 140–210 bp. The optimal range was expected to optimally increase TFx in tumor samples while maintaining stable TFx in controls, or to result in a significantly higher increase of TFx in tumor than in control samples. CNV profiles were visualized using R package ggplot2 v3.4.2[21], displaying genome-wide copy number variations as log2 ratios. Recurrent CNVs were identified in plasma and urine samples with detectable ctDNA and an estimated tumorfraction >10%. Frequencies of copy number states (neutral, loss, gain) were calculated across 1000 kb bins and visualized with frequencies scaled from 0 to 1, where 1 indicated that all analyzed samples harbored a gain or a loss in a specific genomic region.

Additionally, chromosomal instability analysis (CIA) was performed based on lcWGS data from plasma and urinary cfDNA and a synoptic CIA score was calculated, following previously published approaches[37,38] with minor adaptations. For each tumor or control sample, genome-wide read counts normalized for GC content and mappability without further (PoN) correction were obtained for 2601 × 1000 kb windows, as previously computed with the ichorCNA algorithm. Then, absolute z-scores for each 1000 kb bin were calculated by comparing normalized read counts to the PoN mean, accounting for standard deviations. The CIA score was finally derived from the sum of absolute z-scores in the range of 95–99$^{th}$ percentiles, providing a quantitative measure of chromosomal instability across tumor and control samples.

*Fresh-frozen tissue and matched buffy coat samples (gDNA)*

CNV profiling was also performed on lcWGS data from eight PCa tissue and matched buffy coat samples, with buffy coat data serving as a copy-neutral reference (PoN) for each tissue sample. CNVs in buffy coat samples were also assessed using individualized PoNs, excluding the analyzed sample from the reference set. The CNV workflow was similar to the one from the LBx analysis, but excluded in-silico size selection since sheared gDNA was used. The ichorCNA algorithm was applied with default parameters, except for custom adjustments in the HMM settings for 1x bulk tumors (default: 0.1x cfDNA). All chromosomes were analyzed, but only autosomes were used for parameter estimation. Since all patients and controls were male, gender was fixed as "male". For buffy coat samples, normal contamination settings were adjusted to: --normal "c(0.9, 0.95, 0.99, 0.995, 0.999, 1)". TFx was estimated for all PCa tissue and buffy coat samples. CNV profiles were visualized, displaying genome-wide copy number distributions (log2 ratios), and recurrent alterations in CNV-positive tissue samples were identified, as described for LBx samples.

Fragmentation analysis of plasma and urinary cfDNA

Plasma and urinary cfDNA fragment distributions were analyzed by assessing insert sizes from lcWGS data. Insert sizes were extracted from aligned, filtered, and deduplicated BAM files using Samtools v1.15.1[12] (samtools view -f66 file.bam | cut -f 9 > insert_sizes.txt). Relative frequencies were calculated as the proportion of each size within this range, and cumulative frequencies were also derived. Insert sizes were restricted to 30–700 bp fragment lengths. Relative and cumulative fragment length distributions were visualized with R package ggplot2 v3.4.2[21] for individual samples and as median frequencies of tumor and control groups. The Kolmogorov-Smirnov test compared median cumulative distributions between tumor patients and controls. Statistical parameters derived from relative frequency distributions included median and mean fragment length, maximum relative frequency, modal fragment length, cumulative frequency at the modal fragment length, and relative/cumulative frequencies at theoretical mono- (167 bp) and di-nucleosomal (334 bp) peaks. Proportions of predefined fragment length ranges were computed and statistically compared between groups. Fragmentation analysis was adapted from Mouliere et al. [35,39,40], with assessed fragment length ranges including P30–60, P30–100, P30–150, P30–180, P90–150, P160–180, P163–169, P180–220, P150–300, P250–320, P250–420, P324–344, and P420–700 bp. Ratios of short-tolong fragments and nucleosome-associated ranges were analyzed, such as P30–150 bp/P160–180 bp, P90–150 bp/P163–169 bp, P30–150 bp/P150–300 bp, P30–150 bp/P163–169 bp, P30–100 bp/P160–180 bp, P30–100 bp/P163–169 bp and P160–180 bp/P250–420 bp. Additionally, the 10bp-oscillation pattern was examined in 30–150 bp and 150–300 bp fragment length ranges, characterized by alternating maxima and minima with a distance of approximately 10 bp between each two maxima/minima. Local maxima and minima were identified within 5 bp sliding windows. The maxima heights and minima depths were determined from the relative frequency values. An oscillation score was calculated as the sum of local maxima heights minus minima depths in both the 30–150 bp and 150–300 bp fragment length ranges.

Detection of circulating tumor-derived DNA (ctDNA) in plasma and urine

CtDNA detection in plasma and urine from lPCa and advanced PCa (aPCa) patients was assessed using cfDNA fragmentation features (10bp-oscillation score for plasma, P163–169 bp for urine), estimated TFx from CNV analysis, CIA score, and methylation score. TFx, CIA score, and methylation score were evaluated in both plasma and urinary cfDNA, whereas two different features were used for plasma (10bp-oscillation score) and urinary (P163-169bp) cfDNA fragmentation. TFx, CIA score, methylation score and P163–169 bp in urinary cfDNA used a ctDNA detectability threshold set at the 95$^{th}$ percentile of control samples; tumor samples with values above this threshold were considered positive. The 10bp-oscillation score in plasma cfDNA (30–150 bp fragment length) was lower in tumor samples compared to controls, and the ctDNA detectability threshold was set at the 5$^{th}$ percentile of controls, classifying tumor samples below this as positive.

### Supplementary Note

Validation of PCa-specific methylation markers with a published LBx methylation dataset

Robustness and performance of the 67 hypermethylated DMRs identified in PCa tissue and validated using the published dataset from Börno et al.[23] were assessed with the application to another published LBx methylation dataset from PCa patients[24] (**Supplementary Figure S3**). Chen et al. performed cfMeDIP-seq on 133 plasma samples, including 30 samples from lPCa patients and 103 samples from mCRPC patients, collected at baseline, during treatment, or at progression. Sequencing results from cancer-free controls were not available in the external data set. Assessment of absolute methylation levels within the 67 selected regions revealed a clear distinction between lPCa and mCRPC, in both PCA and hierarchical clustering (**Supplementary Figure S3**). To assess specificity of the selected regions, PCA was also performed on randomly selected 67 regions with similar CpG density distribution (CpG density >6), showing weaker separation between lPCa and mCRPC (**Supplementary Figure S3**). The external dataset comprised two distinct cohorts — primary lPCa and mCRPC — for which genome-wide molecular differences were expected, though potential confounding factors or batch effects could not be excluded. To broaden the comparative framework, hierarchical clustering of β-values in the 67 tissue-informed, hypermethylated regions was performed on plasma samples from the external dataset (Chen et al.[24]), combined with plasma or urine samples from our cohort (**Supplementary Figure S4**). In both LBx sources, most cancer-free controls and lPCa/aPCa patients from our cohort clustered with lPCa patients from the external dataset. These samples generally showed low β-values, while mCRPC patients from the external cohort exhibited high β-values across most regions. Two plasma samples from aPCa patients in our cohort clustered with mCRPC samples from the external cohort, showing distinct methylation signals and also harbored detectable ctDNA in the genomic analysis (increased TFx and CIA score) which confirmed a strong tumor signal.

Low methylation signals in both the external cohort of lPCa patients and most PCa patients in our cohort might indicate very low ctDNA content or insufficient coverage of altered methylation events by the hypermethylated PCa biomarkers. Regional hypermethylation is known to increase with tumor progression[41,42], along with expected ctDNA levels, as reflected by the elevated and heterogeneous methylation signals in mCRPC plasma samples. Chen et al. similarly reported a strong correlation between hypermethylated DMRs and ctDNA content in mCRPC plasma, but they also noted that some mCRPC patients with low or undetectable ctDNA clustered closer to lPCa patients[24]. In our cohort, several plasma samples exhibited increased and variable methylation levels, suggesting the presence of ctDNA and potential tumor aggressiveness. Hierarchical clustering of plasma samples from the external cohort was also supplemented with urine samples from our cohort. This aimed to compare methylation signals between LBx sources, despite targeting the same biomarker regions. Urine samples mostly harbored low β-values, except for some regions in which increased levels were visible. Two urine-specific clusters emerged: one near the mCRPC (plasma) cluster with shared patterns, and another on the heatmap's left side with increased β-values in selected regions. Notably, one urine sample from an aPCa patient clustered with mCRPC plasma samples and was previously identified as a strong outlier with detectable ctDNA (elevated TFx) in our CNV analysis. However, this urine sample did not match the two plasma outliers in our cohort. Additionally, eight urine samples from two lPCa and six aPCa patients clustered with the mCRPC group, but at the outermost edge of the cluster. Overall, urinary cfDNA from lPCa and aPCa patients exhibited similar methylation patterns to plasma, but with generally higher levels in biomarker regions, suggesting a better representation of PCa-specific methylation biomarkers. This might reflect a higher ctDNA content in urine, possibly due to direct shedding from the urinary tract[43,44], or other biological and technical differences between LBx sources.

<u>Genome-wide assessment of CNVs in PCa tissue samples</u>

CNV analysis in PCa tissue identified samples with high, low, or no detectable genomic alterations (**Supplementary Figure S6**). Five tissue samples exhibited CNVs, with four showing predominantly deletions and one displaying widespread genomic instability with both deletions and amplifications. The sample with both amplifications and deletions corresponded to an aggressive Gleason 9 tumor (pT3b) with lymph node metastases (pN1 (2/22)). The other four CNV-positive samples had a Gleason 7a pattern, with two tumors confined to the prostate (pT2c, N0/Nx) and two locally advanced cases (pT3a–b, one with pN1). The remaining three samples, despite sufficient tumor cell content (40–90%) and large tumor sizes (2× pT3a, 1× pT3b), showed no detectable CNVs. Comparisons between CNV-positive tissue samples and matched plasma and urine samples were inconclusive, as all LBx samples had TFx below the ctDNA detectability threshold. These findings are supported by previous studies describing CNV-based molecular subgroups in primary PCa[45,46]. The Cancer Genome Atlas Research Network identified three categories: highly unstable genomes with detectable CNVs, CNV-free tumors, and an intermediate group comprising ~50% of cases[45]. Taylor et al. reported six CNV clusters, with four showing minimal alterations, predominantly deletions, or nearly unaltered genomes and two exhibiting high CNV burden, including genome-wide amplifications and deletions or mainly gains in 8q and chromosome 7[46]. Our results align with these classifications, as the eight PCa tissue samples could be assigned to either unaltered, deletion-dominant, or high CNV burden groups, with the Gleason 9 tumor falling into the latter. The observed alterations also matched common genomic changes in primary PCa, including deletions in 8p, 13q, and chromosome 18, as well as in 5p, 6p, and 12p[45,47] (**Supplementary Figure S6**). The tissue sample with detectable amplifications showed known gains in chromosomes 7 and 8q[45,47], alongside with additional alterations on 3, 5, 11, 12, and 13 (**Supplementary Figure S6**).

<u>Assessment of 10bp-oscillation patterns in plasma and urinary cfDNA fragmentation</u>

The cfDNA fragmentation analysis included an assessment of the 10bp-oscillation pattern in the 30–150 bp fragment length range, characterized by periodic local maxima and minima, which is thought to arise from nucleosome positioning and periodic enzymatic cleavage of DNA[48,49]. A synoptic oscillation score was calculated based on amplitude differences to evaluate these characteristic fragmentation patterns. Previous studies by Mouliere et al. demonstrated that this score differs significantly between tumor and control samples, making it a potential biomarker for tumor characterization[35,39]. In our cohort, plasma cfDNA harbored local maxima at 51, 61, 71, 81, 92, 102, 112, 122, 133, and 144 bp, while local minima followed 3–5 bp later at 54, 66, 75, 86, 97, 107, 116, 127, 136, and 145 bp. Tumor patients exhibited a significantly reduced 10bp-oscillation amplitude in plasma cfDNA compared to controls. The oscillation score progressively decreased from controls to lPCa to aPCa, with notably lower scores in tumor patients ($p = 0.02$, adjusted $p = 0.17$), including differences between lPCa and controls ($p = 0.048$, adjusted $p = 0.07$) and between aPCa and controls ($p = 0.028$, adjusted $p = 0.07$). These findings align with similar trends observed by Mouliere et al. in cerebrospinal fluid from glioma patients, where individuals with detectable CNVs exhibited differences compared to those without CNVs[39]. Given the significant differences observed in plasma cfDNA fragmentation and the supporting literature, the 10bp-oscillation score (30–150 bp) was selected as a key plasma cfDNA fragmentation feature for tumor characterization and comparison with other (epi)genomic features. In urinary cfDNA, a distinct 10bp-oscillation pattern was equally observed, and the proportion of urinary cfDNA fragments within 30–150 bp was slightly higher in PCa patients (0.421) compared to controls (0.403). Local maxima were located at 41, 52, 62, 72, 83, 93, 103, 114, 124, 133, 142, and 148 bp, while local minima appeared slightly shifted to the right at 45, 56, 66, 77, 87, 97, 108, 118, 128, 137, and 146 bp. The highest oscillation score was seen in lPCa, decreased in controls and further in aPCa (**Supplementary Figure S10**). However, due to high variability within cohorts, no significant differences were observed.

Beyond the 30–150 bp range, our data revealed an extended oscillation pattern in fragments with 150–300 bp length, particularly pronounced in urinary cfDNA (**Supplementary Figure S10**). While the plasma cfDNA oscillation pattern remained irregular with inconsistent numbers of local maxima and minima, urinary cfDNA exhibited a structured 10 bp-periodicity with up to 15 additional oscillations. Local maxima were found at 159, 175, 196, 210, 220, 228, 237, 242, 250, 258, 267, 275, 283, and 291 (293) bp, while local minima were located at 165, 183, 201, 208, 216, 225, 233, 241, 248, 257, 266, 274, 283, 291, and 297 bp. The proportion of 150–300 bp fragments was higher in tumors, with the highest levels seen in aPCa. However, no significant differences were observed between groups (**Supplementary Figure S10**). Chandrananda et al. reported three additional peaks at 151, 173, and 177 bp with reduced ~5 bp periodicity in longer plasma cfDNA fragments[48], while a detailed characterization of the 150–300 bp oscillation pattern in urinary cfDNA has not been previously documented.

### Supplementary Figures and Tables

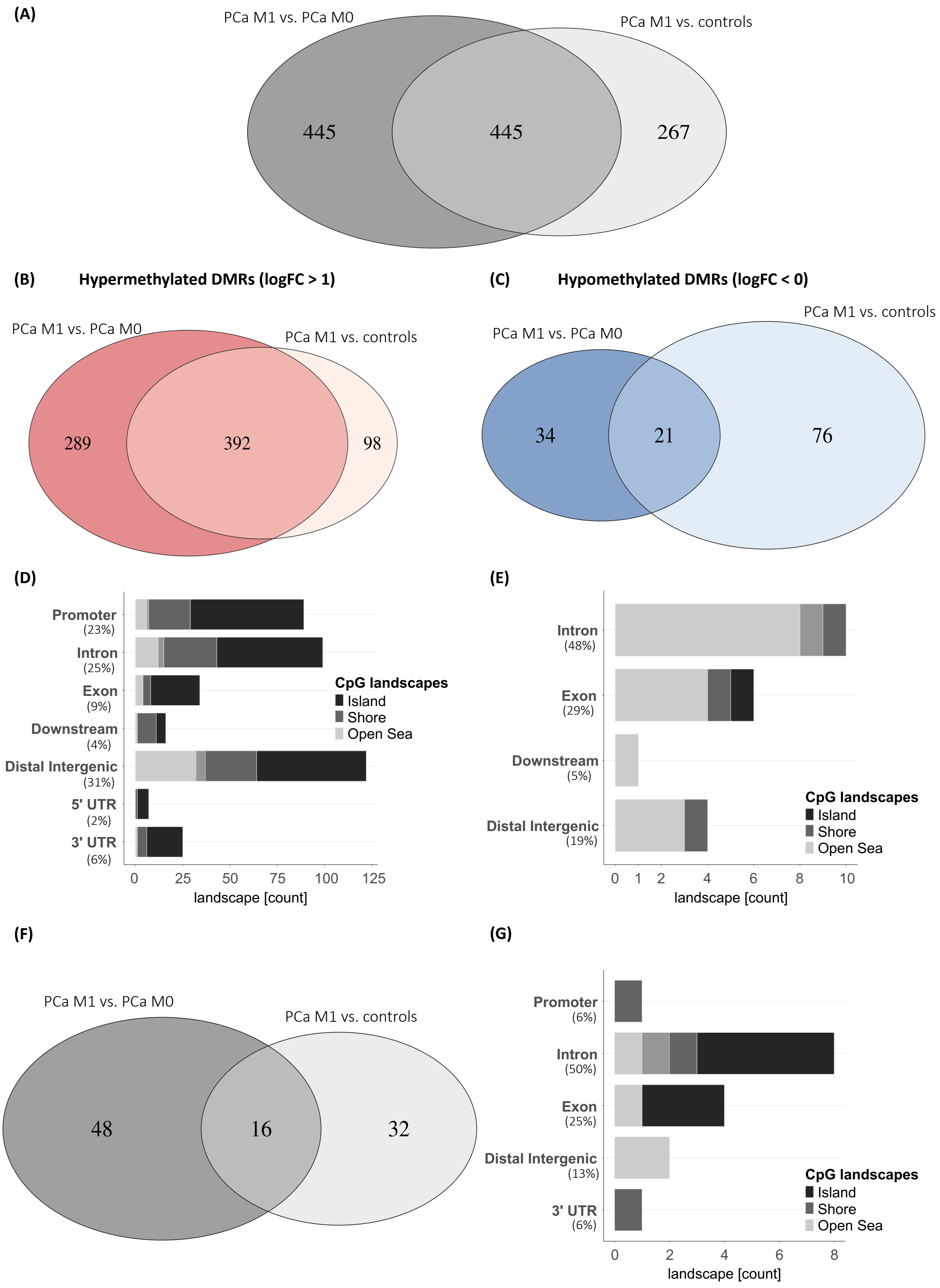


**Supplementary Figure S1:** Genome-wide methylation profiling in plasma and urinary cfDNA and determination of DMRs between PCa patients and cancer-free controls. (A) Common significant DMRs in plasma cfDNA between the comparisons metastatic PCa patients (PCa M1) vs. controls or PCa M1 vs. PCa patients without distant metastases (PCa M0). (B) Common significant, hypermethylated DMRs (logFC > 1) in plasma cfDNA between the comparisons PCa M1 vs. controls or PCa M1 vs. PCa M0. (C) Common significant, hypomethylated DMRs (logFC < 0) in plasma cfDNA between the comparisons PCa M1 vs. controls or PCa M1 vs. PCa M0. (D) Genomic annotation (location within 3’ or 5’UTR, distal intergenic region, downstream region, exon or intron, promotor region) and assessment of CpG-associated landscapes (island, open sea, shelf, shore) within common significant, hypermethylated DMRs (logFC > 1) in plasma cfDNA between the comparisons PCa M1 vs. controls or PCa M1 vs. PCa M0. (E) Genomic annotation (location within 3’ or 5’UTR, distal intergenic region, downstream region, exon or intron, promotor region) and assessment of CpG-associated landscapes (island, open sea, shelf, shore) within common significant, hypomethylated DMRs (logFC < 0) in plasma cfDNA between the comparisons PCa M1 vs. controls or PCa M1 vs. PCa M0. (F) Common significant DMRs in urinary cfDNA between the comparisons PCa M1 vs. controls or PCa M1 vs. PCa M0. (G) Genomic annotation (location within 3’ or 5’UTR, distal intergenic region, downstream region, exon or intron, promotor region) and assessment of CpG-associated landscapes (island, open sea, shelf, shore) within common significant DMRs in urinary cfDNA between the comparisons PCa M1 vs. controls or PCa M1 vs. PCa M0. cfDNA = cell-free DNA, DMRs = differentially methylated regions, logFC = log fold change, M0/M1 = absence/presence of distant metastases, n = number, PCa = prostate cancer, UTR = untranslated region

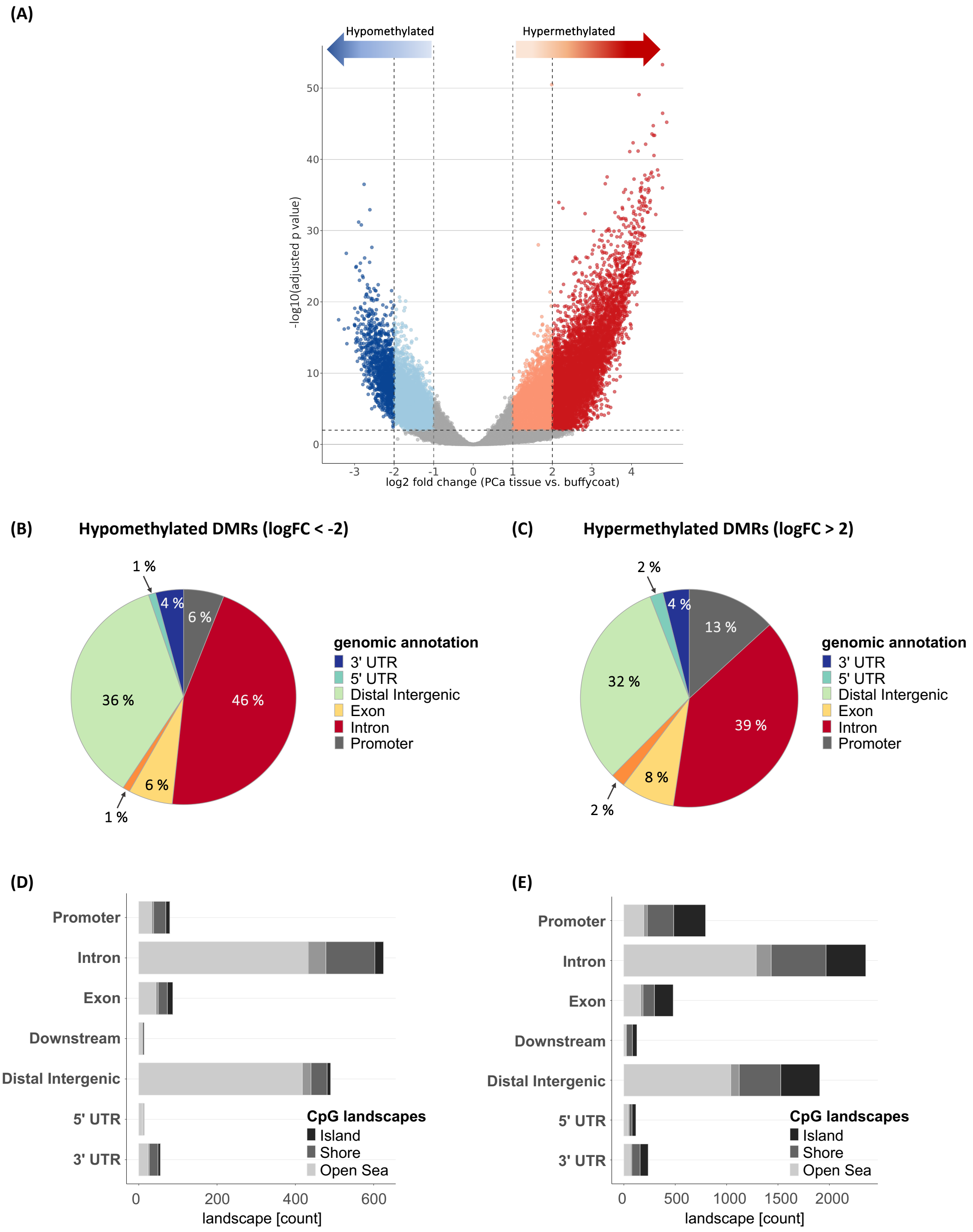


**Supplementary Figure S2:** Genome-wide methylation profiling in genomic DNA from PCa tissue and determination of DMRs between eight PCa tissue and matched buffy coat samples. (A) Overview of the results from the DMR analysis between eight PCa tissue and matched buffy coat samples. Distribution of log2-fold changes and adjusted p values in 810,174 genomic regions. Correction for multiple testing was performed with Benjamini-Hochberg method, significance was defined as adjusted p value < 0.01. Vertical dashed lines: log2FC = +- 1 and +-2, respectively; horizontal dashed line: adjusted p value = 0.01. (B) Genomic annotation (location within 3' or 5'UTR, distal intergenic region, downstream region, exon or intron, promotor region) of 1364 significant, hypomethylated DMRs (logFC < -2) in PCa tissue vs. buffy coat. (C) Genomic annotation (location within 3' or 5'UTR, distal intergenic region, downstream region, exon or intron, promotor region) of 6015 significant, hypermethylated DMRs (logFC > 2) in PCa tissue vs. buffy coat. (D) Assessment of CpG-associated landscapes (island, open sea, shelf, shore) in 1364 significant, hypomethylated DMRs (logFC < -2) in PCa tissue vs. buffy coat. (E) Assessment of CpG-associated landscapes (island, open sea, shelf, shore) in 6015 significant, hypermethylated DMRs (logFC > 2) in PCa tissue vs. buffy coat.

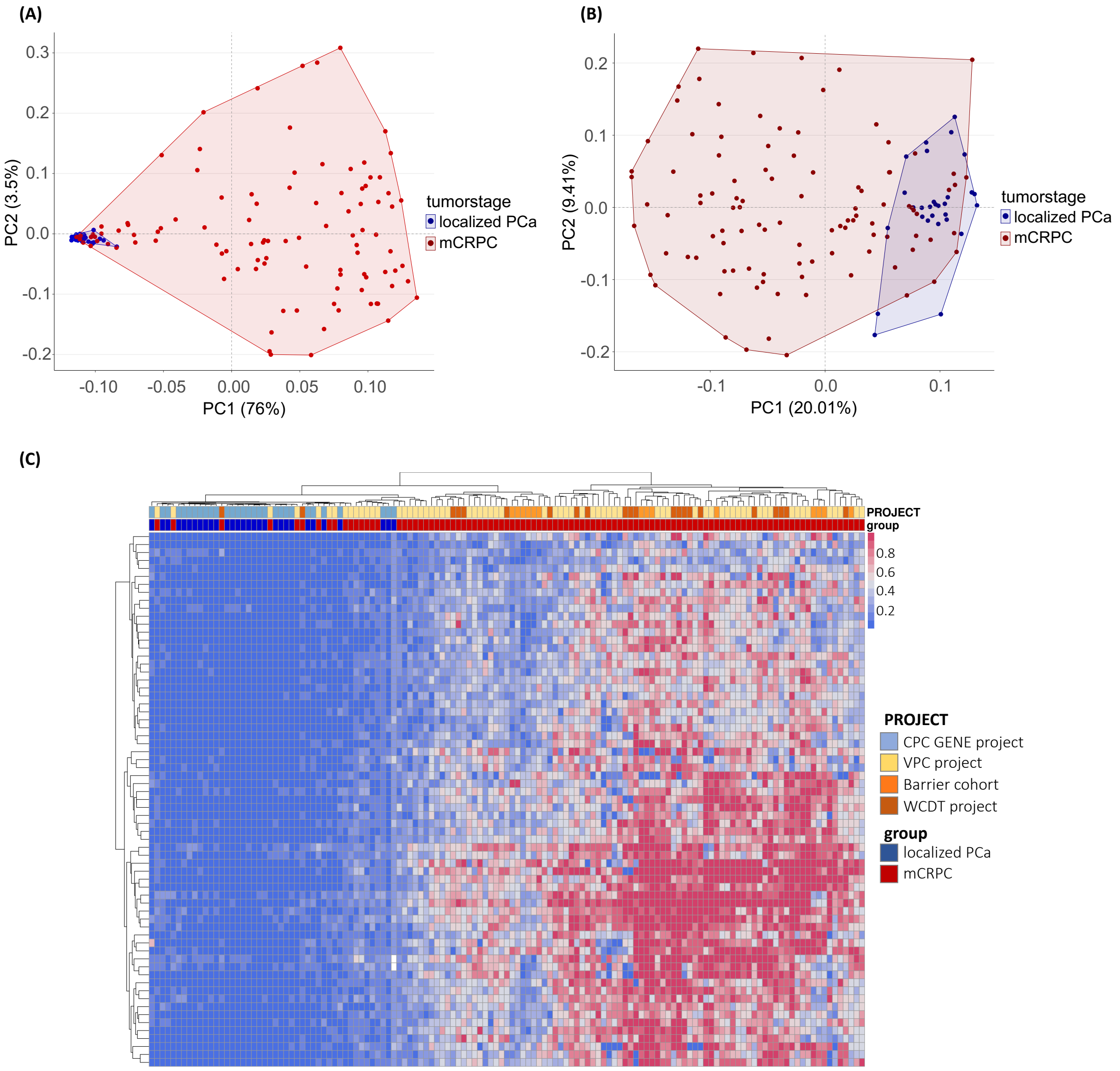


**Supplementary Figure S3:** Clustering based on β-values in 67 marker regions (300 bp regions) for plasma samples from lPCa or mCRPC patients from an external cohort (Toronto, Chen et al. [24]). (A) PCA based on β-values in the 67 marker regions. (B) PCA based on β-values in 67 randomly selected, genome-wide 300bp-regions with similar CpG density compared to the 67 marker regions from own tissue analysis. (A+B) X- and y-axes display the first and second component, explaining the most and second most proportion of the variance, respectively, with the respective proportions (%) reported next to the axis title. (C) Hierarchical clustering based on β-values in the 67 marker regions. The project names were derived from the original publication[24]. lPCa = localized prostate cancer, mCRPC = metastatic castration-resistant prostate cancer, PCA = principal component analysis

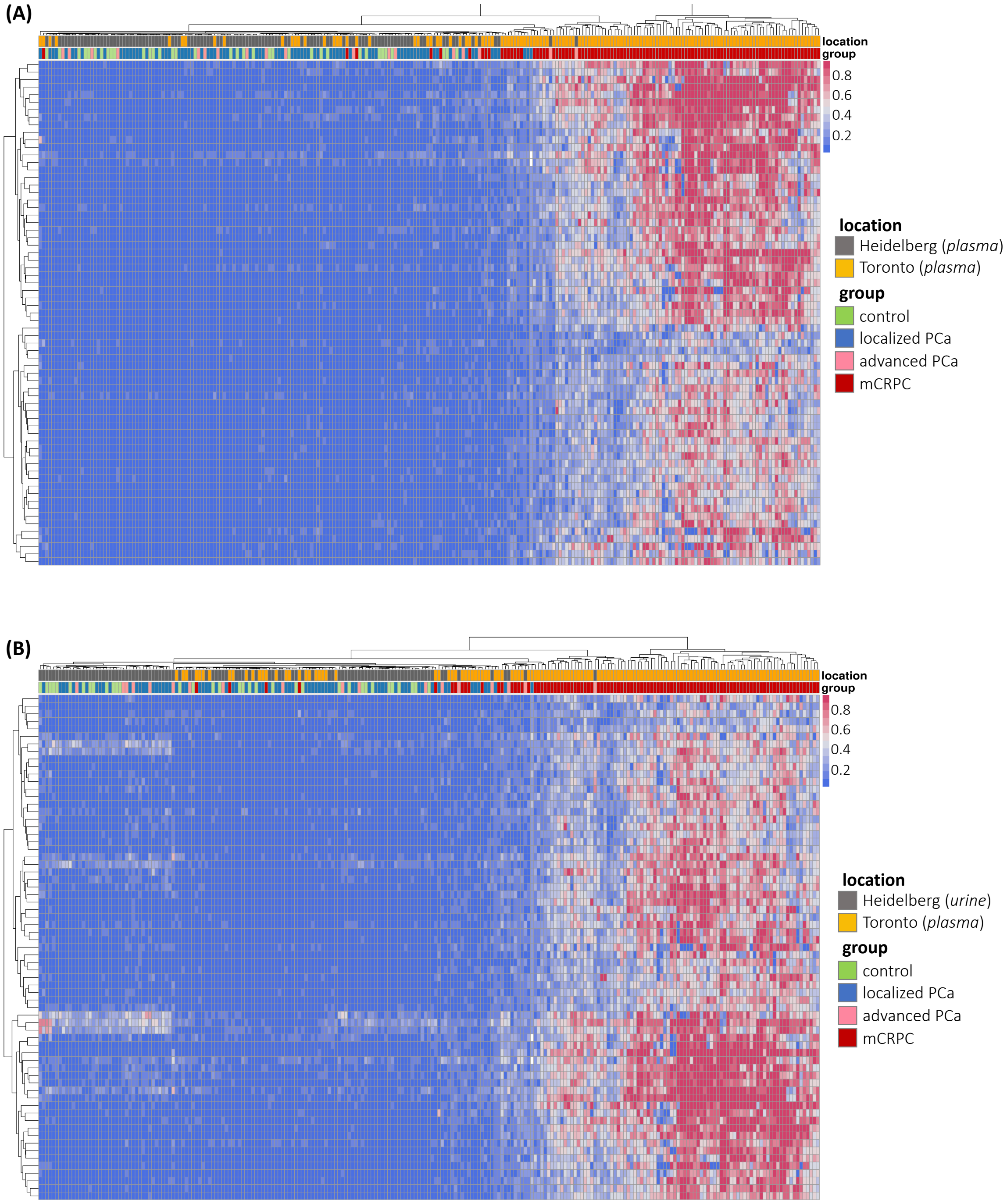


**Supplementary Figure S4:** Hierarchical clustering based on β-values in 67 marker regions (300 bp regions) for plasma samples from lPCa or mCRPC patients from an external cohort (Toronto, Chen et al. [24]), in addition to plasma (A) or urine (B) samples from cancer-free controls, primary lPCa and aPCa patients from the own cohort (Heidelberg). aPCa = advanced prostate cancer

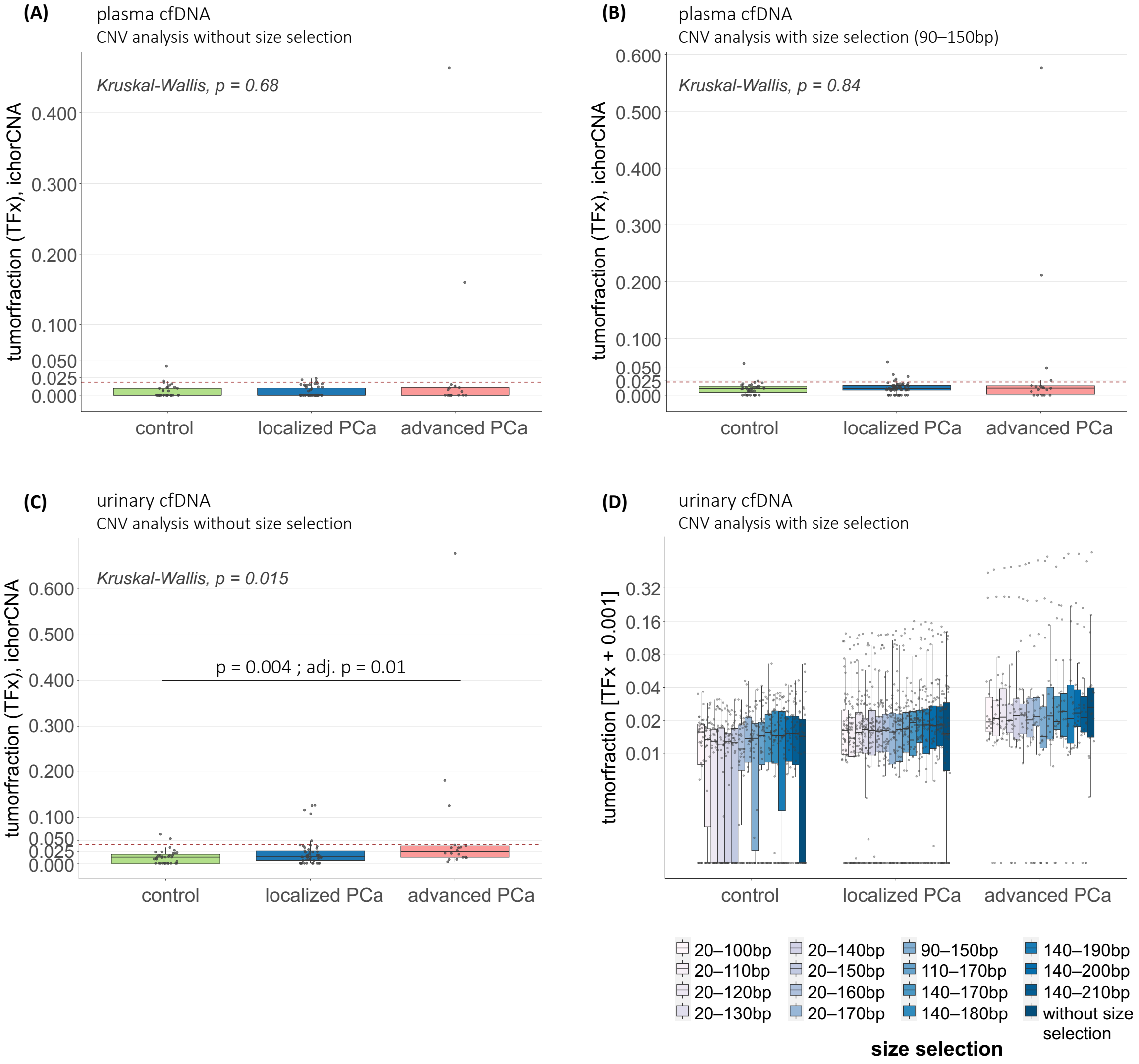


**Supplementary Figure S5:** Genome-wide assessment of CNVs in plasma and urinary cfDNA based lcWGS data. (A) Tumorfractions estimated with the ichorCNA algorithm based on the CNV analysis without size selection in plasma samples from cancer-free controls, lPCa patients, and aPCa patients. (B) Tumorfractions estimated with the ichorCNA algorithm based on the CNV analysis after in-silico size selection for cfDNA fragments with 90–150bp lengths in plasma samples from cancer-free controls, lPCa patients, and aPCa patients. (C) Tumorfractions estimated with the ichorCNA algorithm based on the CNV analysis without size selection in urine samples from cancer-free controls, lPCa patients, and aPCa patients. (A-C) The horizontal, red dotted line represents the ctDNA detectability threshold (95$^{th}$ percentile of control cohort). Results between the three cohorts were statistically compared with Kruskal-Wallis testing, significant results were defined as p value <0.05. (D) Tumorfractions estimated with the ichorCNA algorithm based on the CNV analysis after in-silico size selection for different cfDNA fragment length ranges in urine samples from cancer-free controls, lPCa patients, and aPCa patients. Log2-transformed y-axis; TFx with pseudo count (0.001). (A-D) Box plot center lines indicate the median, and boxes illustrate the interquartile range with Tukey whiskers. Each dot represents one sample. bp = base pairs, CNVs = copy number variations, lcWGS = low-coverage whole-genome sequencing, TFx = tumorfraction

(A)

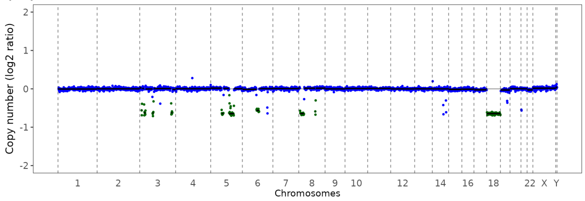


Gleason 7a ; pT2c pNx (cN0 cM0) L0 V0 Pn1;
80% tumor cell content

(B)

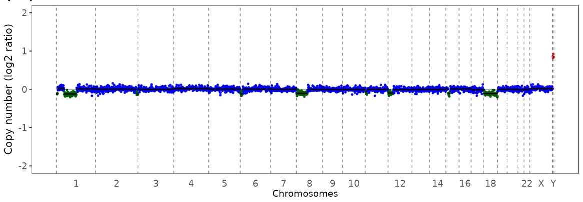


Gleason 7a; pT3b pN1 (1/21) cM0 L1 V0 Pn1;
60% tumor cell content

(C)

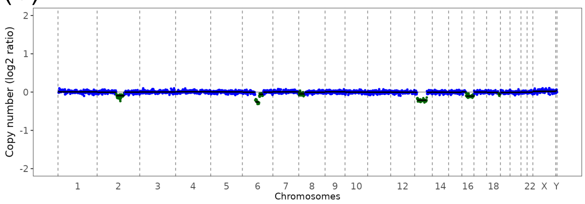


Gleason 7a; pT2c pNx (cN0 cM0) L0 V0 Pn1;
70% tumor cell content

(D)

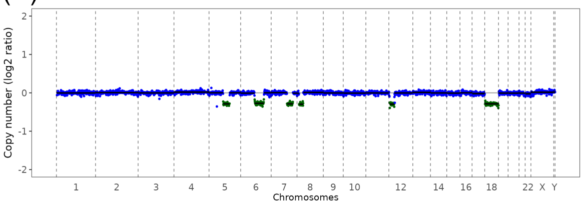


Gleason 7a; pT3a pNx (cN0 cM0) L0 V0 Pn1;
70% tumor cell content

(E)

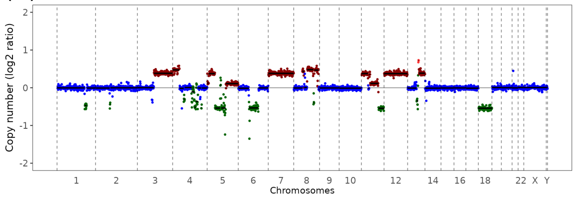


Gleason 9; pT3b pN1 (2/22) cM0 L1 V0 Pn1;
70% tumor cell content

(F)

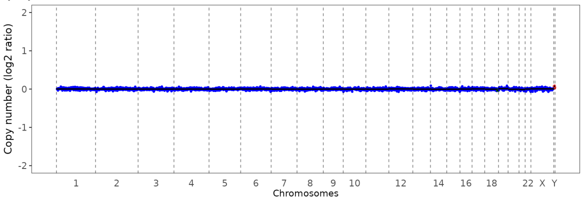


Gleason 7a; pT3b pNx (cN0 cM0) V0 Pn0;
50% tumor cell content

(G)

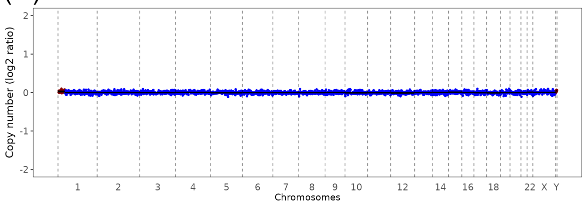


Gleason 7a; pT3a pN0 (0/8) cM0 L1 V0 Pn1;
90% tumor cell content

(H)

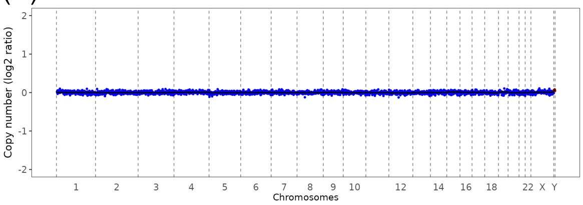


Gleason 7a; pT3a pNx (cN0 cM0) L0 V0 Pn1;
40% tumor cell content

(I)

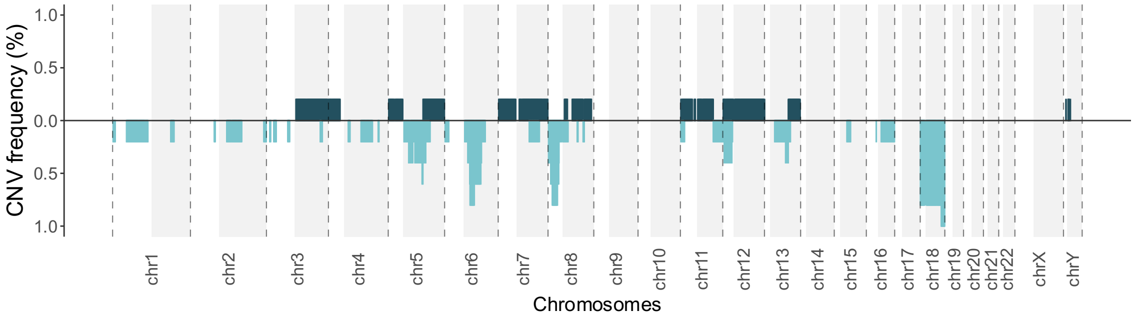


**Supplementary Figure S6:** Genome-wide assessment of CNVs in genomic DNA from fresh-frozen PCa tissue samples based on lcWGS data. (A–H) Single CNV profiles of eight PCa tissue samples. (A–E) PCa tissue samples with detectable alterations. (F–H) PCa tissue samples without detectable alterations. (I) Summary of recurrent amplifications and deletions in 5 PCa tissue samples with detectable CNVs and TFx >10%. The y-axis indicates the frequency of a detected copy number state at the chromosomal coordinate specified on the x-axis across the samples. Areas shaded in gray represent the q-arm of the respective chromosome. chr = chromosome, cM = clinical assessment of distant metastases, cN/pN = clinical/pathological assessment of lymph node metastases, L0/L1 = absence/presence of lymphatic invasion, N0/N1 = absence/presence of lymph node metastases, Nx = unknown lymph node status, V0 = no venous invasion, Pn0/Pn1 = absence/presence of perineural invasion, pT = pathological assessment of the primary tumor's extent

**(A)**

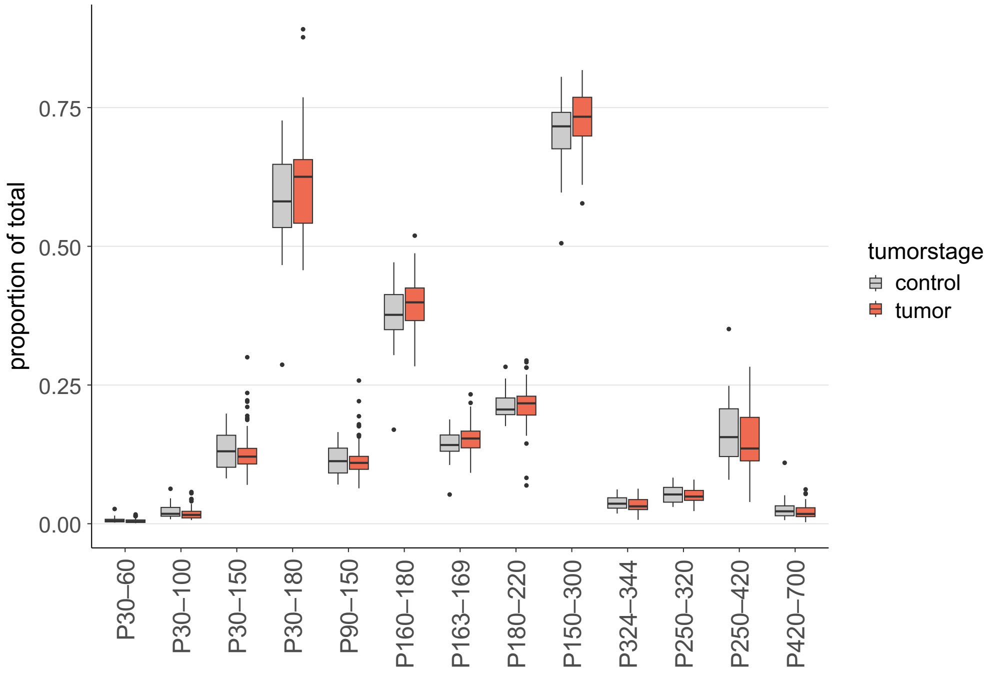


**(B)**

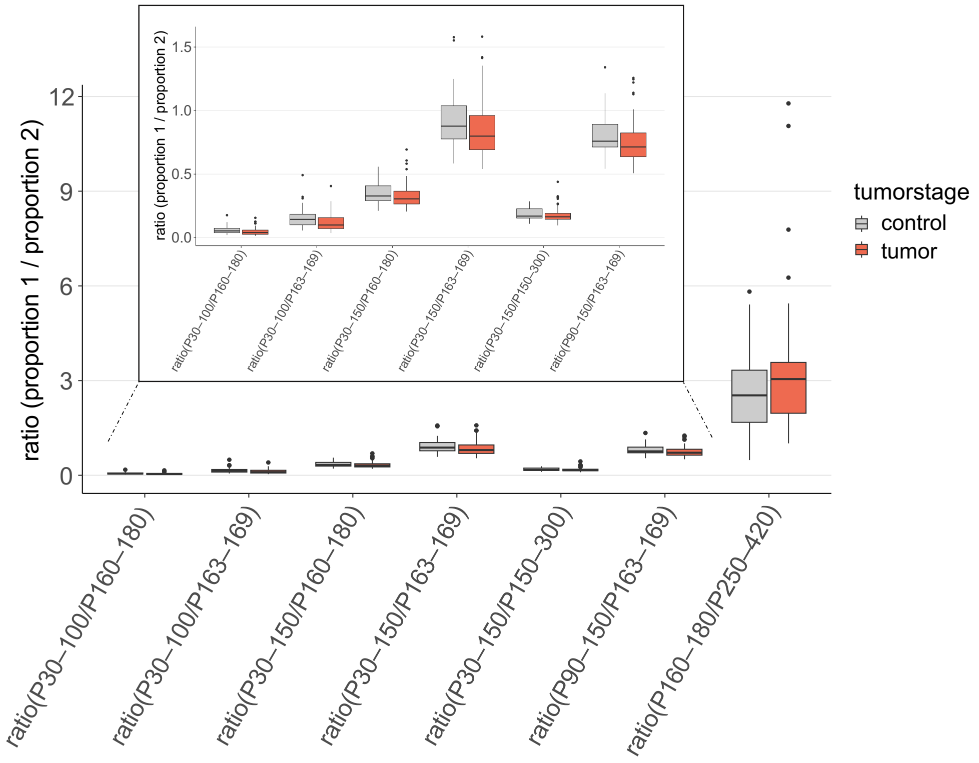


**Supplementary Figure S7:** Plasma cfDNA fragmentation analysis. (A) Different proportions of plasma cfDNA fragment length ranges (in relation to all fragments with 30–700 bp length) in all tumor and control samples. (B) Different ratios of proportions of plasma cfDNA fragment length ranges (in relation to all fragments with 30–700 bp length) in all tumor and control samples. The additional box displays a zoomed window. (A+B) Box plot center lines indicate the median, and boxes illustrate the interquartile range with Tukey whiskers. Dots represent outlying samples.

(A)

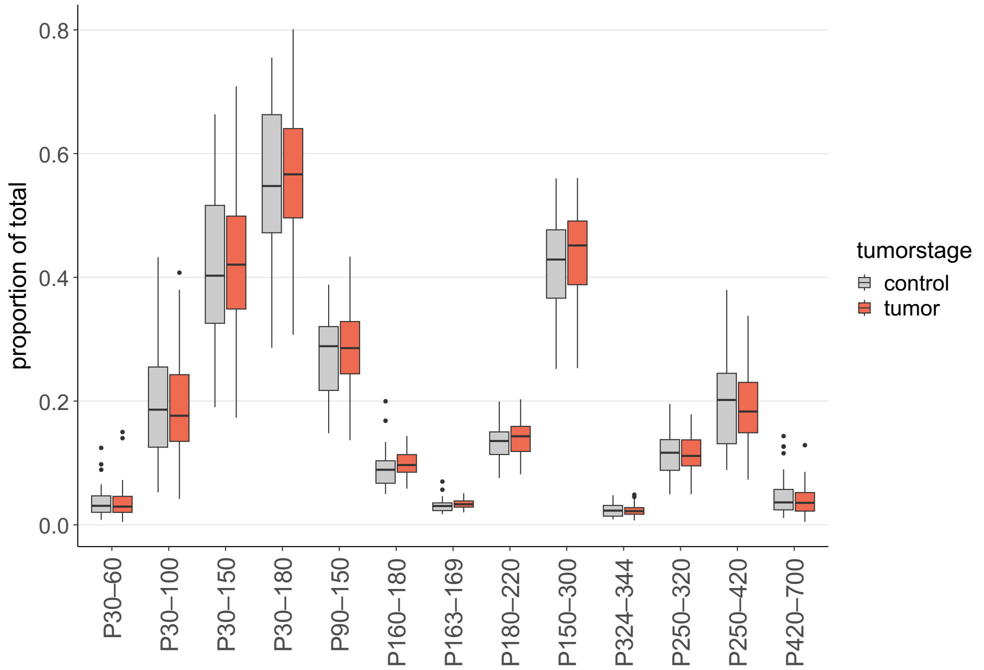


(B)

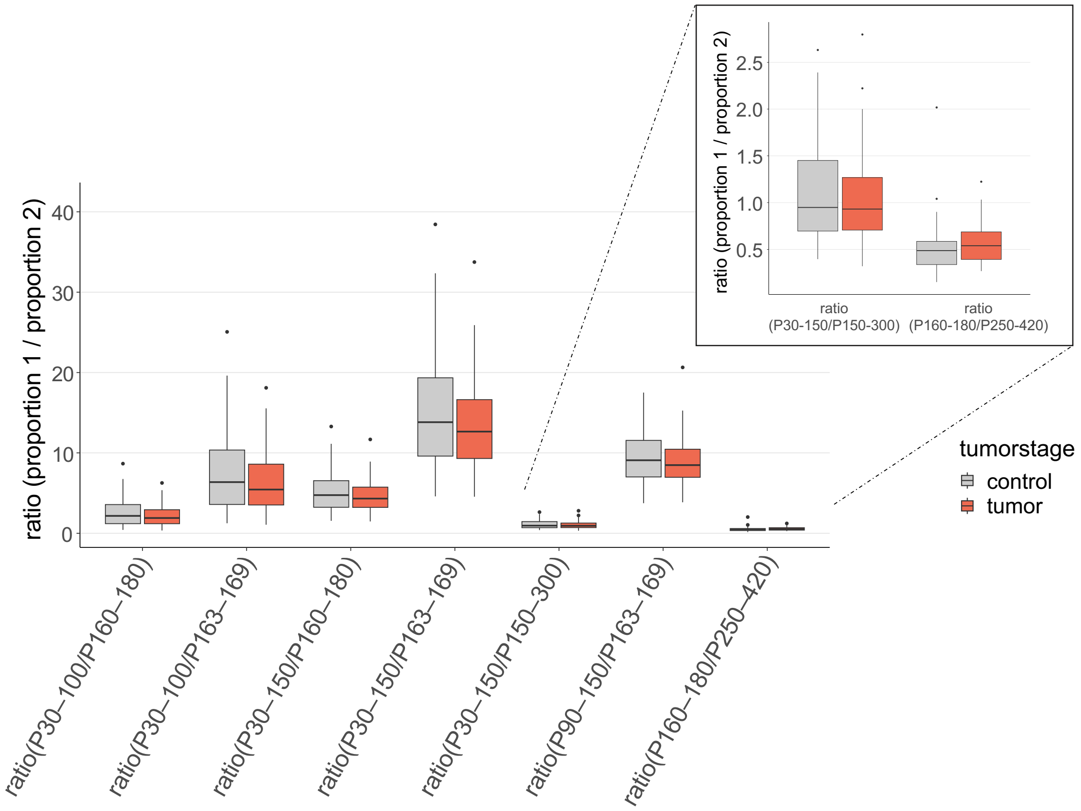


**Supplementary Figure S8:** Urinary cfDNA fragmentation analysis. (A) Different proportions of urinary cfDNA fragment length ranges (in relation to all fragments with 30–700 bp length) in all tumor and control samples. (B) Different ratios of proportions of urinary cfDNA fragment length ranges (in relation to all fragments with 30–700 bp length) in all tumor and control samples. The additional box displays a zoomed window. (A+B) Box plot center lines indicate the median, and boxes illustrate the interquartile range with Tukey whiskers. Dots represent outlying samples.

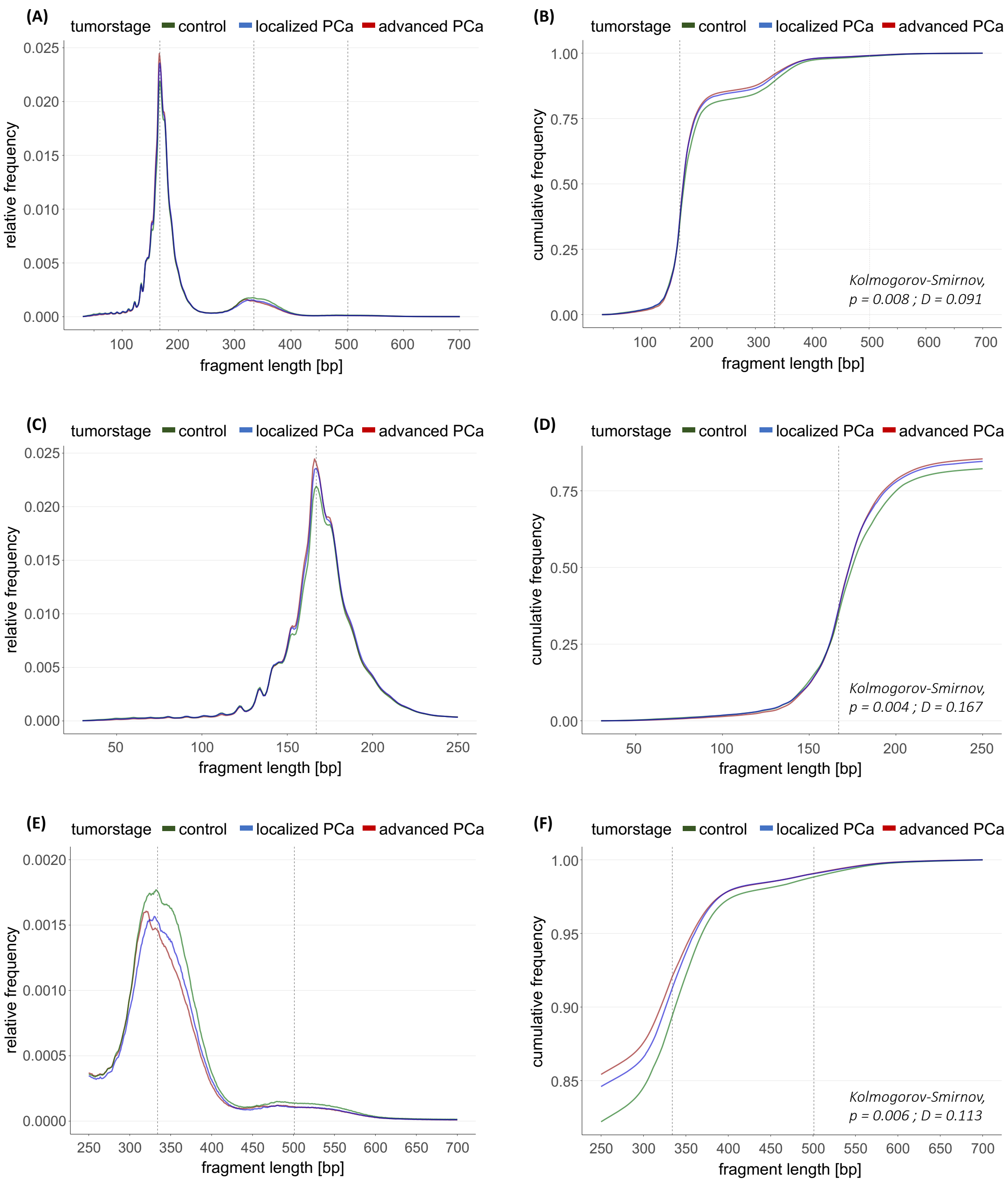


**Supplementary Figure S9:** Relative and cumulative frequency distributions of plasma cfDNA fragmentation derived from lcWGS data. (A–F) Plasma cfDNA fragmentation profiles represented as median profiles of all samples from lPCa and aPCa patients, and cancer-free controls. (A+B) Distribution within fragment length range 30–700 bp. (C+D) Distribution within fragment length range 30–250 bp. (E+F) Distribution within fragment length range 250–700 bp. Y-axis: (A, C, E) relative frequencies of cfDNA fragments with specific length (bp) compared to all fragments (30–700 bp fragment length) and (B, D, F) cumulative frequencies. Vertical dotted grey line(s) indicate 167 bp and its multiples, 334 bp (2 x 167 bp) and 501 bp (3 x 167 bp). Median cumulative distributions between all tumor samples and control samples were compared with Kolmogorov-Smirnov testing. D = distance (D statistic)

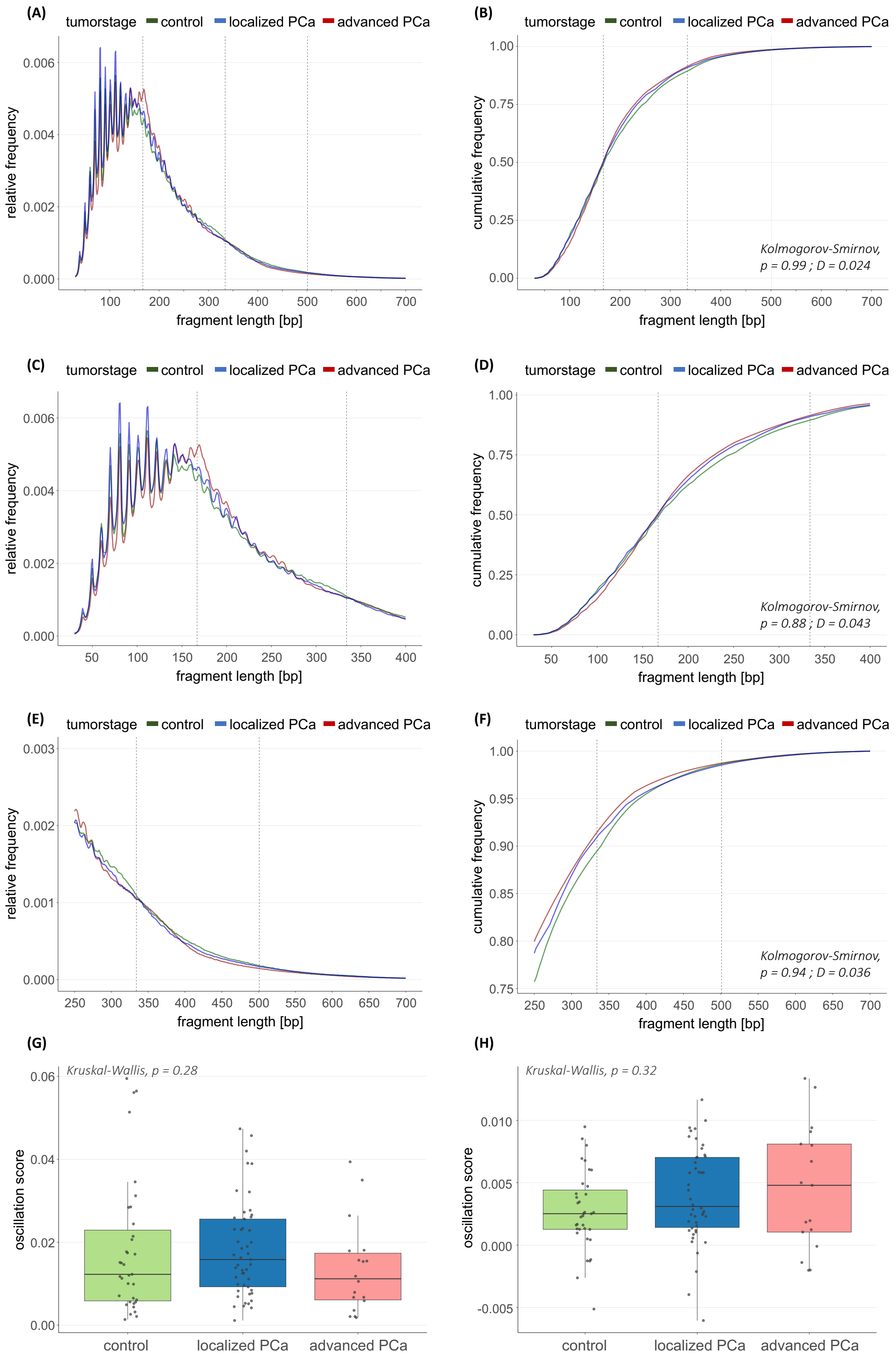


**Supplementary Figure S10:** Relative and cumulative frequency distributions of urinary cfDNA fragmentation derived from lcWGS data. (A–F) Urinary cfDNA fragmentation profiles represented as median profiles of all samples from lPCa and aPCa patients, and cancer-free controls. (A+B) Distribution within fragment length range 30–700 bp. (C+D) Distribution within fragment length range 30–400 bp. (E+F) Distribution within fragment length range 250–700 bp. Y-axis: (A, C, E) relative frequencies of cfDNA fragments with specific length (bp) compared to all fragments (30–700 bp fragment length) and (B, D, F) cumulative frequencies. Vertical dotted grey line(s) indicate 167 bp and its multiples, 334 bp (2 x 167 bp) and 501 bp (3 x 167 bp). Median cumulative distributions between all tumor samples and control samples were compared with Kolmogorov-Smirnov testing. (G) 10bp-oscillation scores in urinary cfDNA fragments with 30–150 bp fragment length from cancer-free controls, lPCa patients, and aPCa patients. (H) 10bp-oscillation scores in urinary cfDNA fragments with 150–300 bp fragment length from cancer-free controls, lPCa patients, and aPCa patients. (G+H) Box plot center lines indicate the median, and boxes illustrate the interquartile range with Tukey whiskers. Each dot represents one sample. Results between the three cohorts were statistically compared with Kruskal-Wallis testing, significant results were defined as p value < 0.05.

**Supplementary Table S1:** Overview of raw, filtered and final sequencing reads based on lcWGS and (cf)MeDIP-seq data. cfDNA = cell-free DNA, (cf)MeDIP-seq = (cell-free) methylated DNA immunoprecipitation sequencing, lcWGS = low-coverage whole-genome sequencing, M = Million

| | **total raw reads**, M | **total final reads**, M | **final reads**, % | **total filtered reads**, M | **filtered reads**, % |
|---|---|---|---|---|---|
| cfDNA, lcWGS | 91.0 [45.4 ; 141.2 ] | 72.4 [35.5 ; 109.8] | 80.28 [70.16 ; 84.56] | 18.1 [9.8 ; 31.4 ] | 19.72 [15.45 ; 29.84] |
| plasma, lcWGS | 88.2 [56.3 ; 117.0 | 71.2 [45.3 ; 95.2] | 81.32 [72.54 ; 84.56] | 16.9 [9.9 ; 23.6 ] | 18.69 [15.45 ; 27.46] |
| urine, lcWGS | 92.3 [45.4 ; 141.2 ] | 73.2 [35.5 ; 109.8] | 79.51 [70.16 ; 83.36] | 19.2 [9.8 ; 31.4 ] | 20.5 [16.64 ; 29.84] |
| cfDNA, cfMeDIP | 90.1 [74,626 ; 218.9 ] | 60.2 [53,358 ; 137.2] | 66.78 [38.55 ; 76.01] | 30.7 [21,268 ; 81.7 ] | 33.22 [24.0 ; 61.45] |
| plasma, cfMeDIP | 91.6 [70.2 ; 200.5] | 62.4 [36.2 ; 136.4] | 68.24 [50.82 ; 76.01] | 29.0 [20.2 ; 64.0 ] | 31.76 [24.0 ; 49.18] |
| urine, cfMeDIP | 88.4 [74,626 ; 218.9 ] | 56.2 [53,358 ; 137.2 ] | 64.04 [38.55 ; 75.45] | 31.8 [21,268 ; 81.7 ] | 35.96 [24.55 ; 61.45] |
| genomic DNA, lcWGS | 76.6 [68.2 ; 87.9] | 62.4 [55.8 ; 71.2 ] | 81.76 [80.78 ; 82.59] | 14.0 [12.2 ; 16.7] | 18.24 [17.41 ; 19.22] |
| tissue, lcWGS | 78.4 [68.2 ; 83.7 ] | 63.5 [55.8 ; 68.0 ] | 81.43 [80.78 ; 82.01] | 14.7 [12.5 ; 16.0 ] | 18.58 [17.99 ; 19.22] |
| buffy coat, lcWGS | 73.3 [70.1 ; 87.9 ] | 60.1 [57.9 ; 71.2 ] | 82.07 [81 ; 82.59] | 13.3 [12.2 ; 16.7 ] | 17.93 [17.41 ; 19.0] |
| genomic DNA, MeDIP | 74.4 [49.7 ; 108.0 ] | 54.1 [33.9 ; 80.0 ] | 72.38 [67.44 ; 74.73] | 20.5 [15.8 ; 28.1 ] | 27.62 [25.27 ; 32.56] |
| tissue, MeDIP | 73.3 [49.7 ; 80.2 ] | 52.6 [33.9 ; 57.7 ] | 70.58 [67.44 ; 74.73] | 20.7 [15.8 ; 26.1 ] | 29.42 [25.27 ; 32.56] |
| buffy coat, MeDIP | 74.7 [68.3 ; 108.0 ] | 54.3 [50.3 ; 80.0 ] | 73.02 [72.07 ; 74.02] | 20.5 [17.8 ; 28.1 ] | 26.98 [25.98 ; 27.93] |

**Supplementary Table S2:** Overview of detectable DMRs between PCa patients and controls in plasma and urinary cfDNA. DMR analysis based on β-values in 300 bp windows, including only windows in which at least one sample harbors >2 NRPKMs, multiple testing with Benjamini-Hochberg (adjusted p value < 0.05). DMR = differentially methylated region, hyper = hypermethylated, hypo = hypomethylated, M0/M1 = absence/presence of distant metastases, N0/N1 = absence/presence of lymph node metastases, NRPKM = normalized reads per kilobase million mapped reads, # = number of

| **comparison** | **# samples** total (cohort 1 vs. cohort 2) | **PLASMA** # significant windows (DMRs) | **URINE** # significant windows (DMRs) |
|---|---|---|---|
| **PCa (all)** vs. **all controls** | plasma: 109 (73 vs. 36)<br>urine: 102 (67 vs. 35) | 0 / 1,168,518 windows | 0 / 1,195,872 windows |
| **aPCa (M1)** vs. **all controls** | plasma: 45 (9 vs. 36)<br>urine: 44 (9 vs. 35) | 712 / 980,927 windows<br>*hyper: 615*<br>*hypo.: 97* | 48 / 992,774 windows<br>*hyper: 29*<br>*hypo.: 19* |
| **aPCa (all)** vs. **all controls** | plasma: 54 (18 vs. 36)<br>urine: 53 (18 vs. 35) | 0 / 1,008,208 windows | 2 / 1,030,695 windows |
| **lPCa high-risk** vs. **all controls** | plasma: 45 (9 vs. 36)<br>urine: 44 (9 vs. 35) | 0 / 945,422 windows | 0 / 980,210 windows |
| **lPCa intermediate-risk** vs. **all controls** | plasma: 78 (42 vs. 36)<br>urine: 72 (37 vs. 35) | 0 / 1,112,508 windows | 0 / 1,127,546 windows |
| **lPCa (all)** vs. **all controls** | plasma: 91 (55 vs. 36)<br>urine: 84 (49 vs. 35) | 0 / 1,134,888 windows | 0 / 1,152,290 windows |
| **lPCa (all) + aPCa (N1 M0)** vs. **all controls** | plasma: 100 (64 vs. 36)<br>urine: 93 (58 vs. 35) | 0 / 1,147,368 windows | 0 / 1,171,278 windows |
| **aPCa (M1)** vs. **aPCa (N1 M0)** | plasma: 18 (9 vs. 9)<br>urine: 18 (9 vs. 9) | 2 / 838,576 windows | 2 / 821,533 windows |
| **aPCa (all)** vs. **lPCa (all)** | plasma: 73 (18 vs. 55)<br>urine: 67 (18 vs. 49) | 0 / 1,115,932 windows | 0 / 1,112,691 windows |
| **aPCa (M1)** vs. **lPCa (all) + aPCa (N1 M0)** | plasma: 73 (9 vs. 64)<br>urine: 67 (9 vs. 58) | 890 / 1,115,932 windows<br>*hyper: 835*<br>*hypo.: 55* | 64 / 1,112,691 windows<br>*hyper: 44*<br>*hypo.: 20* |
| **aPCa (M1)** vs. **lPCa intermediate-risk** | plasma: 51 (9 vs. 42)<br>urine: 46 (9 vs. 37) | 309 / 1,064,258 windows | 18 / 1,048,296 windows |
| **aPCa (all)** vs. **lPCa intermediate-risk** | plasma: 60 (18 vs. 37)<br>urine: 55 (18 vs. 37) | 0 / 1,086,590 windows | 0 / 1,081,990 windows |
| **aPCa (M1)** vs. **lPCa high-risk** | plasma: 18 (9 vs. 9)<br>urine: 18 (9 vs. 9) | 29 / 830,239 windows | 0 / 806,021 windows |
| **aPCa (all)** vs. **lPCa high-risk** | plasma: 27 (18 vs. 9)<br>urine: 27 (18 vs. 9) | 0 / 895,667 windows | 0 / 889,649 windows |

**Supplementary Table S3:** Overview of positive signals in genomic and epigenomic LBx analyses by PSA range and PCa stage. The table presents the number of lPCa and aPCa patients who showed positive signals in each of the four analysis types, stratified by PSA range. For each PSA range, the number of patients with a positive signal is reported separately for lPCa and aPCa. Additionally, the median PSA level and range for each subgroup are provided. For groups with fewer than three patients, single values were reported instead of the median. CIA = chromosomal instability analysis, LBx = Liquid Biopsy, PCa = prostate cancer, PSA = prostate-specific antigen, TFx = tumor fraction

| analysis type | PSA range | localized PCa | | advanced PCa | |
|---|---|---|---|---|---|
| | | # patients | PSA levels median, range | # patients | PSA levels median, range |
| TFx | < 4 ng/ml | 0 | x | 0 | x |
| | 4-10 ng/ml | 7 | 7.7 [4.3 - 8.1] | 2 | 4.3 ; 7.25 |
| | > 10 ng/ml | 1 | 11 | 3 | 30.9 [11 - 90] |
| CIA score | < 4 ng/ml | 1 | 2.7 | 0 | |
| | 4-10 ng/ml | 7 | 8.1 [4.3 - 9.6] | 3 | 7.1 [4.3 - 7.5] |
| | > 10 ng/ml | 2 | 11 ; 26.4 | 5 | 30.9 [11 - 249] |
| Methylation score | < 4 ng/ml | 0 | x | 0 | x |
| | 4-10 ng/ml | 7 | 8.1 [6.2 - 9.6] | 4 | 5.7 [4.1 - 7.5] |
| | > 10 ng/ml | 2 | 11.6 ; 21.4 | 6 | 25.1 [11 - 249] |
| cfDNA fragmentation | < 4 ng/ml | 1 | 3.2 | 0 | x |
| | 4-10 ng/ml | 6 | 7.7 [4.8 - 9.5] | 0 | x |
| | > 10 ng/ml | 2 | 11.5 ; 11.6 | 5 | 19.3 [11 - 249] |

**Supplementary Table S4:** Distribution of positive signals across genomic and epigenomic LBx analyses by PSA range and PCa stage. The table presents the number of positive lPCa and aPCa patients based on the number of positive results across the four analysis types, stratified by PSA range. The first column categorizes patients by the number of positive signals (0,1,2,3,4 out of 4). For each PSA category, the number of lPCa and aPCa patients with positive signal is reported, along with the median PSA level and range for each group. For groups with fewer than three patients, single values were reported instead of the median.

| # positive analyses | PSA range | localized PCa | | advanced PCa | |
|---|---|---|---|---|---|
| | | # patients | PSA levels median, range | # patients | PSA levels median, range |
| **0 out of 4** | < 4 ng/ml | 3 | 3 [2.7 - 3.9] | 0 | x |
| | 4-10 ng/ml | 21 | 6.8 [4.1 - 9.6] | 2 | 7.2, 7.3 |
| | > 10 ng/ml | 8 | 19.56 [12.1 - 40] | 6 | 22 [17 - 33] |
| **1 out of 4** | < 4 ng/ml | 2 | 2.7 ; 3.2 | 0 | x |
| | 4-10 ng/ml | 7 | 6.3 [4.8 - 9.1] | 1 | 4.1 |
| | > 10 ng/ml | 3 | 21.4 [11.5 - 21.4] | 0 | x |
| **2 out of 4** | < 4 ng/ml | 0 | x | 0 | x |
| | 4-10 ng/ml | 8 | 7.9 [4.3 - 9.6] | 1 | 7.1 |
| | > 10 ng/ml | 2 | 11.0 ; 11.6 | 1 | 18.3 |
| **3 out of 4** | < 4 ng/ml | 0 | x | 0 | x |
| | 4-10 ng/ml | 1 | 8.1 | 2 | 4.3 ; 7.5 |
| | > 10 ng/ml | 0 | x | 3 | 90 [19.3 - 249] |
| **4 out of 4** | < 4 ng/ml | 0 | x | 0 | x |
| | 4-10 ng/ml | 0 | x | 0 | x |
| | > 10 ng/ml | 0 | x | 2 | 11 ; 30.9 |

### References

1. Weinreb JC, Barentsz JO, Choyke PL, et al. PI-RADS Prostate Imaging – Reporting and Data System: 2015, Version 2. *Eur Urol*. 2016;69(1):16-40. doi:https://doi.org/10.1016/j.eururo.2015.08.052
2. Görtz M, Radtke JP, Hatiboglu G, et al. The Value of Prostate-specific Antigen Density for Prostate Imaging-Reporting and Data System 3 Lesions on Multiparametric Magnetic Resonance Imaging: A Strategy to Avoid Unnecessary Prostate Biopsies. *Eur Urol Focus*. 2021;7(2):325-331. doi:https://doi.org/10.1016/j.euf.2019.11.012
3. Radtke JP, Schwab C, Wolf MB, et al. Multiparametric Magnetic Resonance Imaging (MRI) and MRI-Transrectal Ultrasound Fusion Biopsy for Index Tumor Detection: Correlation with Radical Prostatectomy Specimen. *Eur Urol*. 2016;70(5):846-853. doi:10.1016/j.eururo.2015.12.052
4. van Leenders G, van der Kwast TH, Grignon DJ, et al. The 2019 International Society of Urological Pathology (ISUP) Consensus Conference on Grading of Prostatic Carcinoma. *Am J Surg Pathol*. 2020;44(8):e87-e99. doi:10.1097/pas.0000000000001497
5. Shen SY, Burgener JM, Bratman SV, De Carvalho DD. Preparation of cfMeDIP-seq libraries for methylome profiling of plasma cell-free DNA. *Nat Protoc*. 2019;14(10):2749-2780. doi:10.1038/s41596-019-0202-2
6. Di Tommaso P, Chatzou M, Floden EW, Barja PP, Palumbo E, Notredame C. Nextflow enables reproducible computational workflows. *Nat Biotechnol*. 2017;35(4):316-319. doi:10.1038/nbt.3820
7. Kurtzer, Gregory M et. al. *Singularity 2.5.2 - Linux application and environment containers for science.* 2018. Sylabs Inc. https://zenodo.org/records/1308868
8. Ewels PA, Peltzer A, Fillinger S, et al. The nf-core framework for community-curated bioinformatics pipelines. *Nat Biotechnol*. 2020;38(3):276-278.doi:10.1038/s41587-020-0439-x
9. Liu D. Algorithms for efficiently collapsing reads with Unique Molecular Identifiers. *PeerJ*. 2019;7:e8275-e8275. doi:10.7717/peerj.8275
10. Martin M. Cutadapt removes adapter sequences from high-throughput sequencing reads. *EMBnet.journal*. 2011;17(1):10-12. doi:10.14806/ej.17.1.200
11. Langmead B, Salzberg SL. Fast gapped-read alignment with Bowtie 2. *Nat Methods*. 2012;9(4):357-359. doi:10.1038/nmeth.1923
12. Li H, Handsaker B, Wysoker A, et al. The Sequence Alignment/Map format and SAMtools. *Bioinformatics*. 2009;25(16):2078-2079. doi:10.1093/bioinformatics/btp352
13. *FastQC: a quality control tool for high throughput sequence data*. Babraham Institute, UK; 2010. https://www.bioinformatics.babraham.ac.uk/projects/fastqc/
14. Ewels P, Magnusson M, Lundin S, Käller M. MultiQC: summarize analysis results for multiple tools and samples in a single report. *Bioinformatics*. 2016;32(19):3047-3048. doi:10.1093/bioinformatics/btw354
15. Lienhard M, Grimm C, Morkel M, Herwig R, Chavez L. MEDIPS: genome-wide differential coverage analysis of sequencing data derived from DNA enrichment experiments. *Bioinformatics*. 2014;30(2):284-6. doi:10.1093/bioinformatics/btt650
16. Chemi F, Pearce SP, Clipson A, et al. cfDNA methylome profiling for detection and subtyping of small cell lung cancers. *Nat Cancer*. 2022;3(10):1260-1270. doi:10.1038/s43018-022-00415-9
17. Lienhard M, Grasse S, Rolff J, et al. QSEA-modelling of genome-wide DNA methylation from sequencing enrichment experiments. *Nucleic Acids Res*. 2017;45(6):e44. doi:10.1093/nar/gkw1193
18. Amemiya HM, Kundaje A, Boyle AP. The ENCODE Blacklist: Identification of Problematic Regions of the Genome. *Sci Rep*. 2019;9(1):9354.doi:10.1038/s41598-019-45839-z

19. Kolde R. Pheatmap: pretty heatmaps. 2018. https://github.com/raivokolde/pheatmap
20. Wang Q, Li M, Wu T, et al. Exploring Epigenomic Datasets by ChIPseeker. *Curr Protoc*. 2022;2(10):e585. doi:https://doi.org/10.1002/cpz1.585
21. Wickham H. ggplot2: Elegant Graphics for Data Analysis. *Springer-Verlag New York*. 2016. https://ggplot2.tidyverse.org
22. Zhu LJ, Gazin C, Lawson ND, et al. ChIPpeakAnno: a Bioconductor package to annotate ChIP-seq and ChIP-chip data. *BMC Bioinformatics*. 2010;11(1):237. doi:10.1186/1471-2105-11-237
23. Börno ST, Fischer A, Kerick M, et al. Genome-wide DNA methylation events in TMPRSS2-ERG fusion-negative prostate cancers implicate an EZH2-dependent mechanism with miR-26a hypermethylation. *Cancer Discov*. 2012;2(11):1024-35. doi:10.1158/2159-8290.CD-12-0041
24. Chen S, Petricca J, Ye W, et al. The cell-free DNA methylome captures distinctions between localized and metastatic prostate tumors. *Nat Commun*. 2022;13(1):6467. doi:10.1038/s41467-022-34012-2
25. Fraser M, Sabelnykova VY, Yamaguchi TN, et al. Genomic hallmarks of localized, non-indolent prostate cancer. *Nature*. 2017;541(7637):359-364. doi:10.1038/nature20788
26. Annala M, Vandekerkhove G, Khalaf D, et al. Circulating Tumor DNA Genomics Correlate with Resistance to Abiraterone and Enzalutamide in Prostate Cancer. *Cancer Discov*. 2018;8(4):444-457. doi:10.1158/2159-8290.Cd-17-0937
27. Zhao SG, Chen WS, Li H, et al. The DNA methylation landscape of advanced prostate cancer. *Nat Genet*. 2020;52(8):778-789. doi:10.1038/s41588-020-0648-8
28. Broad Institute. Picard Toolkit. 2019. https://broadinstitute.github.io/picard/
29. R Foundation for Statistical Computing. R: A Language and Environment for Statistical Computing. 2020. https://www.R-project.org/
30. Sievert C. Interactive Web-Based Data Visualization with R, plotly, and shiny. *Chapman and Hall/CRC*. 2019. https://plotly-r.com
31. Tang YaH, Masaaki and Li, Wenxuan. ggfortify: Unified Interface to Visualize Statistical Results of Popular R Packages. *The R Journal*. 2016;8(2):474-485.
32. Adalsteinsson VA, Ha G, Freeman SS, et al. Scalable whole-exome sequencing of cell-free DNA reveals high concordance with metastatic tumors. *Nat Commun*. 2017;8(1):1324. doi:10.1038/s41467-017-00965-y
33. Lai D. HMM Copy Utils. 2011. https://github.com/shahcompbio/hmmcopy_utils
34. Lai D, Ha G, Shah S. HMMcopy: Copy number prediction with correction for GC and mappability bias for HTS data. 2021. https://git.bioconductor.org/packages/HMMcopy
35. Mouliere F, Chandrananda D, Piskorz AM, et al. Enhanced detection of circulating tumor DNA by fragment size analysis. *Sci Transl Med*. 2018;10(466). doi:10.1126/scitranslmed.aat4921
36. Smith CG, Moser T, Mouliere F, et al. Comprehensive characterization of cell-free tumor DNA in plasma and urine of patients with renal tumors. *Genome Med*. 2020;12(1):23. doi:10.1186/s13073-020-00723-8
37. Chen Z, Zhang C, Zhang M, et al. Chromosomal instability of circulating tumor DNA reflect therapeutic responses in advanced gastric cancer. *Cell Death Dis*. 2019;10(10):697. doi:10.1038/s41419-019-1907-4
38. Liu H, He W, Wang B, et al. MALBAC-based chromosomal imbalance analysis: a novel technique enabling effective non-invasive diagnosis and monitoring of bladder cancer. *BMC Cancer*. 2018;18(1):659. doi:10.1186/s12885-018-4571-7
39. Mouliere F, Mair R, Chandrananda D, et al. Detection of cell-free DNA fragmentation and copy number alterations in cerebrospinal fluid from glioma patients. *EMBO Mol Med*. 2018;10(12)doi:10.15252/emmm.201809323

40. Mouliere F, Smith CG, Heider K, et al. Fragmentation patterns and personalized sequencing of cell-free DNA in urine and plasma of glioma patients. *EMBO Mol Med*. 2021;n/a(n/a):e12881. doi:https://doi.org/10.15252/emmm.202012881
41. Ehrlich M. DNA Hypomethylation In Cancer Cells. *Epigenomics*. 2009;1(2):239-259. doi:10.2217/epi.09.33
42. Massie CE, Mills IG, Lynch AG. The importance of DNA methylation in prostate cancer development. *J Steroid Biochem Mol Biol*. 2017;166:1-15. doi:10.1016/j.jsbmb.2016.04.009
43. Oshi M, Murthy V, Takahashi H, et al. Urine as a Source of Liquid Biopsy for Cancer. *Cancers (Basel)*. 2021;13(11)doi:10.3390/cancers13112652
44. Tivey A, Church M, Rothwell D, Dive C, Cook N. Circulating tumour DNA — looking beyond the blood. *Nat Rev Clin Oncol*. 2022;19(9):600-612. doi:10.1038/s41571-022-00660-y
45. Abeshouse A, Ahn J, Akbani R, et al. The Molecular Taxonomy of Primary Prostate Cancer. *Cell*. 2015;163(4):1011-1025. doi:https://doi.org/10.1016/j.cell.2015.10.025
46. Taylor BS, Schultz N, Hieronymus H, et al. Integrative genomic profiling of human prostate cancer. *Cancer Cell*. Jul 13 2010;18(1):11-22. doi:10.1016/j.ccr.2010.05.026
47. Abate-Shen C, Shen MM. Molecular genetics of prostate cancer. *Genes Dev*. 2000;14(19):2410-34. doi:10.1101/gad.819500
48. Chandrananda D, Thorne NP, Bahlo M. High-resolution characterization of sequence signatures due to non-random cleavage of cell-free DNA. *BMC Med Genomics*. 2015;8:29. doi:10.1186/s12920-015-0107-z
49. Lo YMD, Chan KCA, Sun H, et al. Maternal Plasma DNA Sequencing Reveals the Genome-Wide Genetic and Mutational Profile of the Fetus. *Sci Transl Med*. 2010;2(61):61ra91-61ra91. doi:doi:10.1126/scitranslmed.3001720
